# Beyond reduced alpha power: a weaker temporal link between alpha and other brain rhythms in Alzheimer’s disease

**DOI:** 10.64898/2026.10.23.26366296

**Authors:** Jinwen Wei, Changsong Zhou

## Abstract

**Background:** Reduced alpha power is among the most consistently reported resting electroencephalography (EEG) features of Alzheimer’s disease (AD). However, alpha activity also interacts with other rhythms over time, a dynamic aspect that power measures do not capture. Whether AD weakens this temporal interaction, and whether such a change carries information beyond reduced alpha power, remains unclear.

**Methods:** We analyzed public eyes-closed resting EEG from 29 healthy controls (HC), 36 patients with AD and 23 with frontotemporal dementia (FTD). After separating periodic from aperiodic activity, we tracked the aperiodic exponent and periodic theta, alpha, beta and gamma activity in consecutive 1-s windows. A vector autoregressive model predicted these five measures from the preceding window. Alpha’s predictive contribution (*G_α_*) was the increase in prediction error for theta, beta and gamma when the model was refitted without alpha as a predictor, with training and testing on separate time blocks of each recording. AD–HC differences were adjusted for age and sex, then for alpha expression (periodic alpha power) and signal quality (dynamic range and spatial reliability), with alpha self-persistence (alpha’s carry-over between windows) as a benchmark. We also tested whether expression and *G_α_* each added information about AD beyond the other. An exploratory Jansen–Rit-family circuit model examined whether expression and *G_α_* can change partly independently.

**Results:** *G_α_* was lower in AD than in HC (adjusted difference in 10³*G_α_*, −3.19; 95% confidence interval −5.25 to −1.13; *p* = 0.0035), with a consistent direction across sensitivity analyses. Adjustment for alpha expression and signal quality retained 75.8% of this difference but only 4.4% of the self-persistence difference. Expression and *G_α_* each added information beyond the other (Holm-adjusted *p* = 0.012 for both). FTD contrasts were inconclusive. In the circuit model, reducing local recurrent excitation lowered alpha expression without an estimated decrease in *G_α_*, whereas reducing downstream input lowered *G_α_* with a smaller change in expression.

**Conclusions:** At the group level, the resting alpha abnormality in AD comprises two partly nonredundant dimensions: reduced expression and a weaker temporal link between alpha and subsequent multi-rhythm activity. Quantifying this temporal link may complement conventional spectral markers of AD.

## Introduction

Changes in resting-state electroencephalography (EEG) and magnetoencephalography (MEG) are among the most consistently reported neurophysiological features of Alzheimer’s disease (AD), with the most prominent involving the alpha rhythm which dominates the eyes-closed resting EEG. In AD, posterior alpha power decreases, the alpha peak shifts to lower frequencies, and theta and delta power increase [1–3]. Furthermore, when oscillatory peaks are separated from the aperiodic background [4], periodic alpha [5, 6] and beta [5, 7] power are reduced, whereas aperiodic findings remain inconsistent [5–8]. Specific alpha measures have also been related to AD pathology: a slower alpha peak frequency to both amyloid and tau [9], lower posterior alpha source activity to higher cerebrospinal fluid tau in prodromal AD [10], and MEG alpha hyposynchrony to tau deposition on positron emission tomography [11]. Alpha changes also follow a longitudinal trajectory that varies with disease stage [12]. Together, these properties — combined with the fact that resting EEG is inexpensive, non-invasive, and widely available — have made alpha measures attractive candidates for patient stratification and monitoring in clinical trials [1, 12, 13]. These changes are not specific to AD; they occur in other neurodegenerative dementias: they are similar or more pronounced in Lewy body disease [14, 15], whereas findings in frontotemporal dementia (FTD) are sparser and less consistent [14, 16]. Alpha measures are also sensitive to medication [17], vigilance [18], eye state [19], and recording procedures [20]. Despite this diversity, all these established measures share a common perspective: they characterize how strongly, and at what frequency, alpha activity is expressed on average across a recording.

A complementary view holds that alpha does not operate in isolation. Alpha activity has been linked to functional inhibition and temporal gating of cortical processing [21, 22] and, together with beta activity, to feedback signaling between cortical areas, whereas gamma activity has been associated with feedforward signaling [23–25]. Consistent with these findings, coupling between rhythms is altered in AD and mild cognitive impairment (MCI), although reported directions of change differ [26–29]. In EEG/MEG studies, alterations have been found in phase–amplitude coupling, in which the amplitude of a faster rhythm varies with the phase of a slower one [26–28], and in bispectral measures of nonlinear phase coupling between frequency bands [29]. Both measures quantify coupling within the same data segment, typically averaged over the recording. The same holds for the functional connectivity measures widely used in EEG studies of AD and MCI, such as coherence between recording sites within a frequency band [30]. By contrast, time-resolved analyses add temporal order: switching between functional connectivity configurations, including cross-frequency ones, has been modeled as a sequence of states [31]. Such sequences show which coupling configuration follows which, but they do not test whether fluctuations of a specific rhythm, such as alpha, are followed by changes in the other rhythms that their own recent activity does not already explain.

This temporal dimension matters. Rhythmic activity can occur in transient bursts rather than as a sustained oscillation [32–34], so how rhythms unfold over seconds carries information that time-averaged measures do not capture. Time-resolved studies have also shown that AD alters how resting brain activity unfolds, as seen in EEG microstates and dynamic network states [35–37]. If alpha activity is functionally linked to other rhythms, this link could be reflected in how its fluctuations relate to the subsequent activity of those rhythms. Directed predictive methods, such as Granger-based and transfer-entropy approaches, address this by testing whether one signal’s past improves prediction of another [38, 39], but in AD they have mainly been applied between recording sites or sources rather than between rhythms [30, 40–43]. One between-rhythm application, in MCI, tested a training effect within patients rather than a difference from healthy aging [44]. To our knowledge, whether AD alters the temporal, directed link from alpha to other rhythms, a quantity we term alpha’s predictive contribution, has therefore received little direct attention.

Answering this question also requires disentangling any such dynamic change from the expression deficit, that is, the reduced alpha power described above. Measurement error can bias a predictor’s estimated contribution: in linear regression, classical error in a single predictor attenuates its coefficient, but when correlated predictors are also measured with error, the bias can go in either direction [45, 46]. Tracking moment-to-moment dynamics requires spectral estimates from short data segments, which are less reliable than estimates from longer recordings [47, 48]. Weaker or more spatially restricted alpha is measured with relatively more error, so an alpha-derived dynamic measure could differ between groups merely as a by-product of reduced expression, in a direction that cannot be assumed in advance. Our research question was therefore twofold: is alpha’s predictive contribution reduced in AD, and does any such reduction carry information beyond reduced alpha expression?

We analyzed a public eyes-closed EEG dataset of 29 healthy controls (HC) and 59 clinically diagnosed patients, 36 with AD and 23 with FTD [49, 50]. After separating periodic from aperiodic spectral activity, we tracked the aperiodic exponent and periodic theta, alpha, beta, and gamma activity in consecutive one-second windows. Next, we used a vector autoregressive model to predict all five in each window from their values in the preceding window. Alpha’s predictive contribution, *G_α_*, was defined as the increase in prediction error for theta, beta, and gamma one second later when alpha in the preceding window (lagged alpha) was removed from this model and the model was refitted (Figure 1). Prediction error was estimated with temporally blocked cross-validation, in which the model is fitted to part of each participant’s recording and tested on a separate time block. *G_α_* is thus an out-of-sample measure of incremental prediction in the Granger sense rather than a direct estimate of anatomical information flow [38, 39]. The primary comparison was AD versus HC, adjusted for age and sex, with FTD contrasts providing clinical context. We then asked whether the AD–HC difference in *G_α_* was explained by alpha expression or by two measures of alpha signal quality, namely its dynamic range and spatial split-half reliability. To benchmark this, we applied the same adjustment to conditional alpha self-persistence, which reflects how strongly alpha activity carries over from one window to the next and is expected to depend closely on alpha signal strength. We also compared profiles across predicting rhythms (sources), predicted rhythms (targets), and scalp regions to test whether the AD–HC difference was specific to alpha, to particular target rhythms, or to posterior regions. Additionally, we tested whether alpha expression and *Gα* each provided group information beyond the other. Finally, we used an exploratory Jansen–Rit-family circuit model [51–53] to ask whether the two measures can change partly independently within a simple generative system. Together, these analyses ask whether the alpha abnormality in AD is better described, at the group level, by two partly nonredundant dimensions than by expression alone: how much alpha is expressed and how much its recent fluctuations contribute to subsequent multi-rhythm activity.

**Figure 1.**
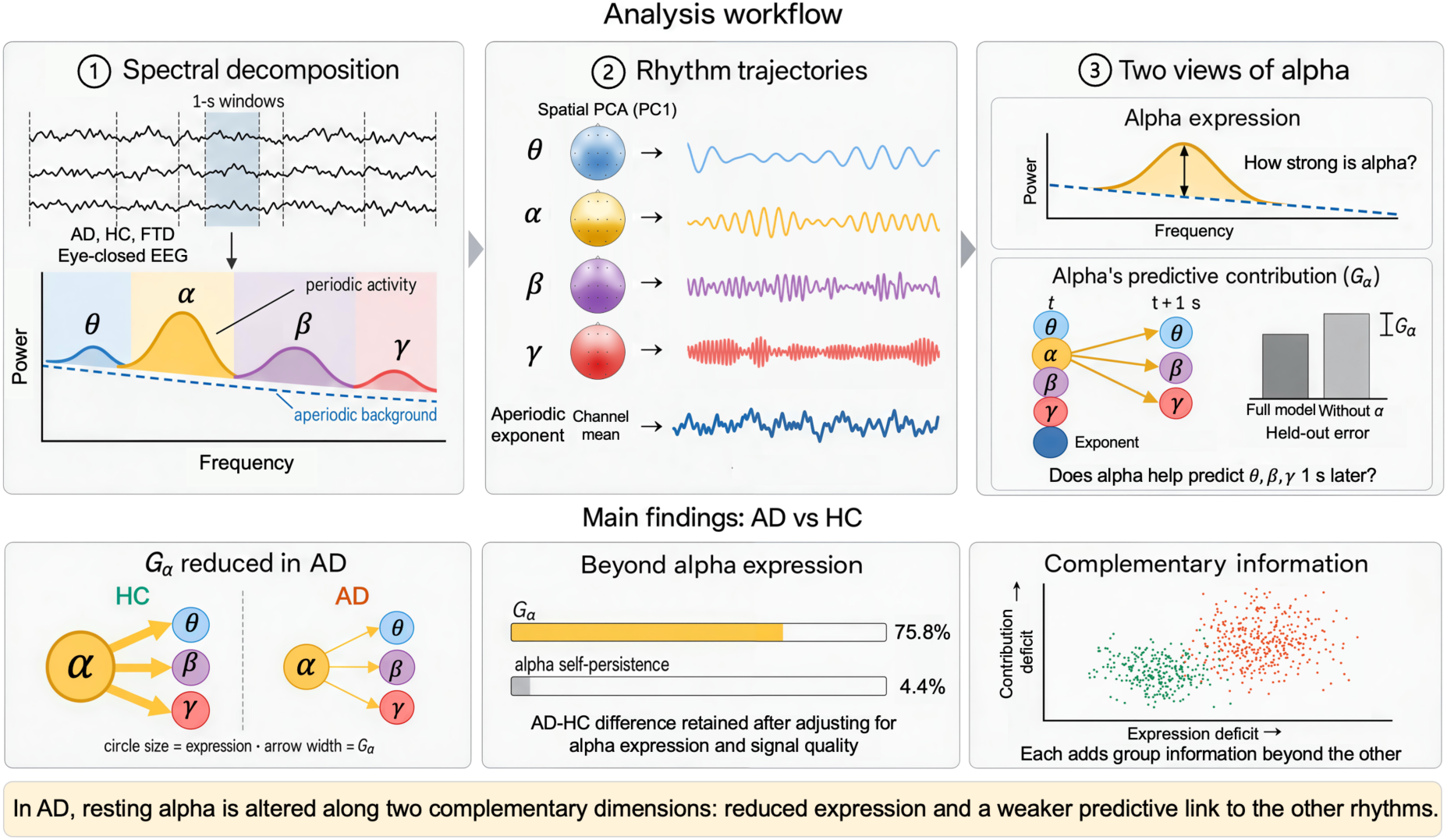
Schematic summary of the analysis workflow and main findings. Top, analysis workflow. (1) Eyes-closed resting EEG from HC, AD and FTD groups was divided into 1-s windows, and periodic activity was separated from the aperiodic background. (2) Band-specific spatial PC1 weights reduced theta, alpha, beta and gamma activity to one trajectory each, and the channel-mean aperiodic exponent formed a fifth. (3) Alpha was characterized by its expression, the periodic alpha power above the background, and by its predictive contribution (*G_α_*), the increase in held-out error for predicting theta, beta and gamma 1 s later when lagged alpha is withheld and the model refitted. **Bottom, main findings.** *G_α_* was lower in AD than in HC. Adjustment for alpha expression and signal quality (alpha dynamic range and spatial split-half reliability) retained 75.8% of this difference but only 4.4% of the corresponding difference in alpha self-persistence, and expression and *G_α_* each added group information beyond the other. Please note that, apart from the retention percentages, all plotted elements are schematic.

## Methods

### Participants and data

We analyzed publicly available, de-identified resting-state EEG recordings from OpenNeuro dataset ds004504 [49, 50]. The cohort comprised 88 participants: 29 healthy controls (HC), 36 participants with AD and 23 with FTD. Diagnoses were made clinically by the source study using established criteria, including the National Institute of Neurological and Communicative Disorders and Stroke–Alzheimer’s Disease and Related Disorders Association (NINCDS-ADRDA) criteria for AD [49]. Diagnostic labels were used as supplied and were not biomarker-confirmed in the present analysis. Cognitive status was assessed with the Mini-Mental State Examination (MMSE). EEG was recorded from 19 scalp electrodes of the international 10–20 system at 500 Hz while participants sat with their eyes closed. We used the preprocessed recordings distributed with the dataset, which had been band-pass filtered at 0.5–45 Hz, re-referenced to the mean of A1 and A2, and cleaned with artifact subspace reconstruction and independent component analysis [49]; no further filtering or re-referencing was applied.

### Spectral decomposition

To follow rhythmic activity from moment to moment, signals were divided into consecutive, non-overlapping 1-s Hann windows (1-Hz resolution). This window length balances signal quality, temporal detail and sample size. Longer windows contain more data and therefore give more reliable spectral estimates [47, 48], but they blur transient changes in rhythmic activity [32–34] and leave fewer windows for fitting the prediction model, whereas shorter windows would lose the 1-Hz frequency resolution used to define the bands. Windows that were incomplete or crossed a recording discontinuity were excluded, and lagged observations never spanned such gaps. Fp1, Fp2, F7 and F8 were excluded from the rhythmic states because of their susceptibility to ocular and frontal artifacts, whereas Fp1 and Fp2 served only as references for removing high-frequency (30–44 Hz) nuisance variation from frontal channels. In each window and channel, the aperiodic background was fitted as a straight line in log–log coordinates over 2–44 Hz, iteratively masking positive residuals so that spectral peaks did not inflate the background, an approach conceptually similar to spectral parameterization [4]. Periodic activity was the mean log-power residual relative to this background in the theta (4–7 Hz), alpha (8–12 Hz), beta (13–29 Hz) and gamma (30–44 Hz) bands (Figure 2A, B). Details are given in Supplementary Methods 1–2.

**Figure 2.**
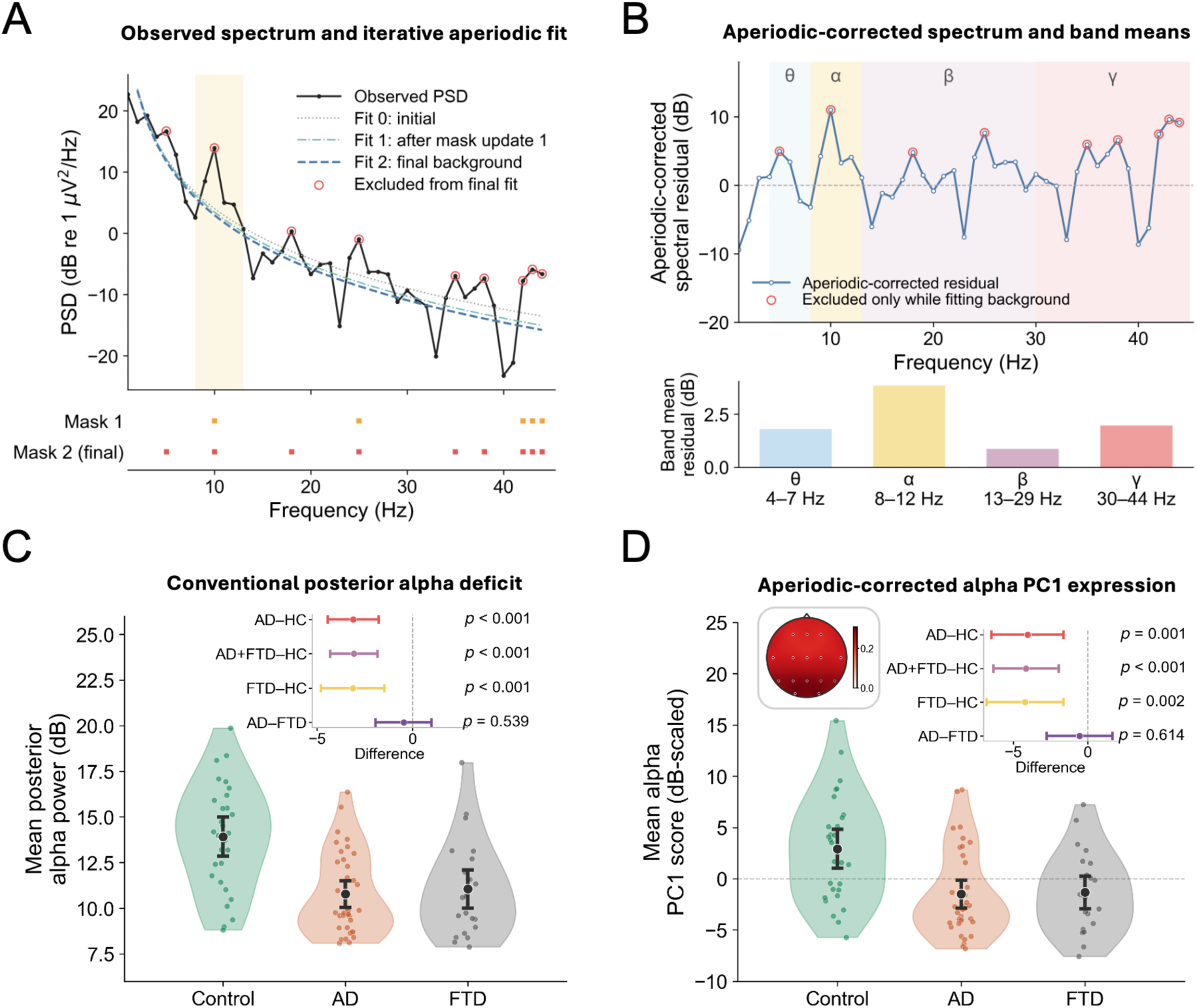
Spectral decomposition and alpha expression. (A) Example spectrum from an illustrative 1-s window showing the initial background fit and two refits. After each of the first two fits, frequency bins with large positive residuals (spectral peaks) were masked so that peaks did not inflate the background; the two rows below the spectrum mark the bins masked at each update. Bins excluded from the final fit (red open circles) were still used to compute the periodic residual in B. The vertical line marks the nominal 45-Hz low-pass boundary. (B) Aperiodic-corrected spectrum for the same window with band means for theta (4–7 Hz), alpha (8–12 Hz), beta (13–29 Hz) and gamma (30–44 Hz); zero denotes equality with the background, and negative values denote power below it. (C) Conventional posterior absolute alpha power (O1, O2, P3, P4 and Pz). (D) PC1-weighted alpha expression; the inset shows the alpha spatial loading. In C and D, points are participants, violins show distributions, and black points and bars show means ± 1.96 standard errors; insets show age- and sex-adjusted contrasts with HC4 95% CIs and wild-bootstrap *p* values.

### Rhythmic state trajectories

To summarize each rhythm by a single, more reliable time course, a fixed spatial weighting was obtained for each band as the first principal component (PC1) of the channel covariance, averaged with equal weight across participants, and each window’s band map was projected onto it. Theta, alpha and beta used 15 channels; gamma used 11 channels, excluding the temporal electrodes T3–T6 to limit muscle contamination. Together with the channel-mean aperiodic exponent, this yielded five state trajectories per participant (the aperiodic exponent and periodic theta, alpha, beta and gamma activity), each standardized within participant (Figure 1). Spatial weights and standardization were fixed before cross-validation, so only model coefficients were estimated within training folds (see details in Supplementary Methods 2).

### Alpha expression and signal quality

To characterize how strongly alpha was expressed and how reliably its time course was captured, we computed the following measures. Alpha expression was the mean periodic alpha power above the aperiodic background across all windows and the 15 channels, expressed in dB. For comparison with previous studies, we also computed conventional posterior absolute alpha power (O1, O2, P3, P4 and Pz, before aperiodic correction) and a PC1-weighted alpha expression score. Two signal-quality measures were computed: alpha dynamic range, the standard deviation of the alpha trajectory before standardization, and spatial split-half reliability, the correlation between two estimates of the same alpha trajectory, each computed from a different half of the 15 channels; high values indicate that alpha fluctuations are measured consistently across channels (see details in Supplementary Methods 3).

### Alpha’s predictive contribution

To quantify how much recent alpha activity helps predict the subsequent activity of the other rhythms, temporal dependencies among the five states were modeled with a first-order vector autoregressive model, VAR(1), in which each state in the next 1-s window was predicted from all five states in the current window, with a fixed ridge penalty (λ = 0.10n, where n is the number of training observations) [54]. Model order was assessed with the Bayesian information criterion (BIC). Within each participant, lagged observations, that is, pairs of consecutive windows, were divided in time order into five contiguous folds. Using contiguous folds rather than randomly selected pairs means that training and test pairs meet only at fold boundaries, which limits how much the similarity of neighboring windows can inflate prediction accuracy [55]. For each held-out fold f, a full model and a reduced model refitted without lagged alpha (alpha in the preceding window) were trained on all window pairs in the remaining four folds, and the increase in held-out mean squared error (MSE) was computed for participant i and target rhythm j and then averaged over the three target rhythms (Figure 3A):

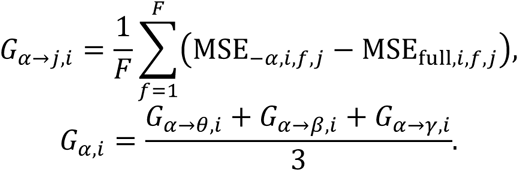

**Figure 3.**
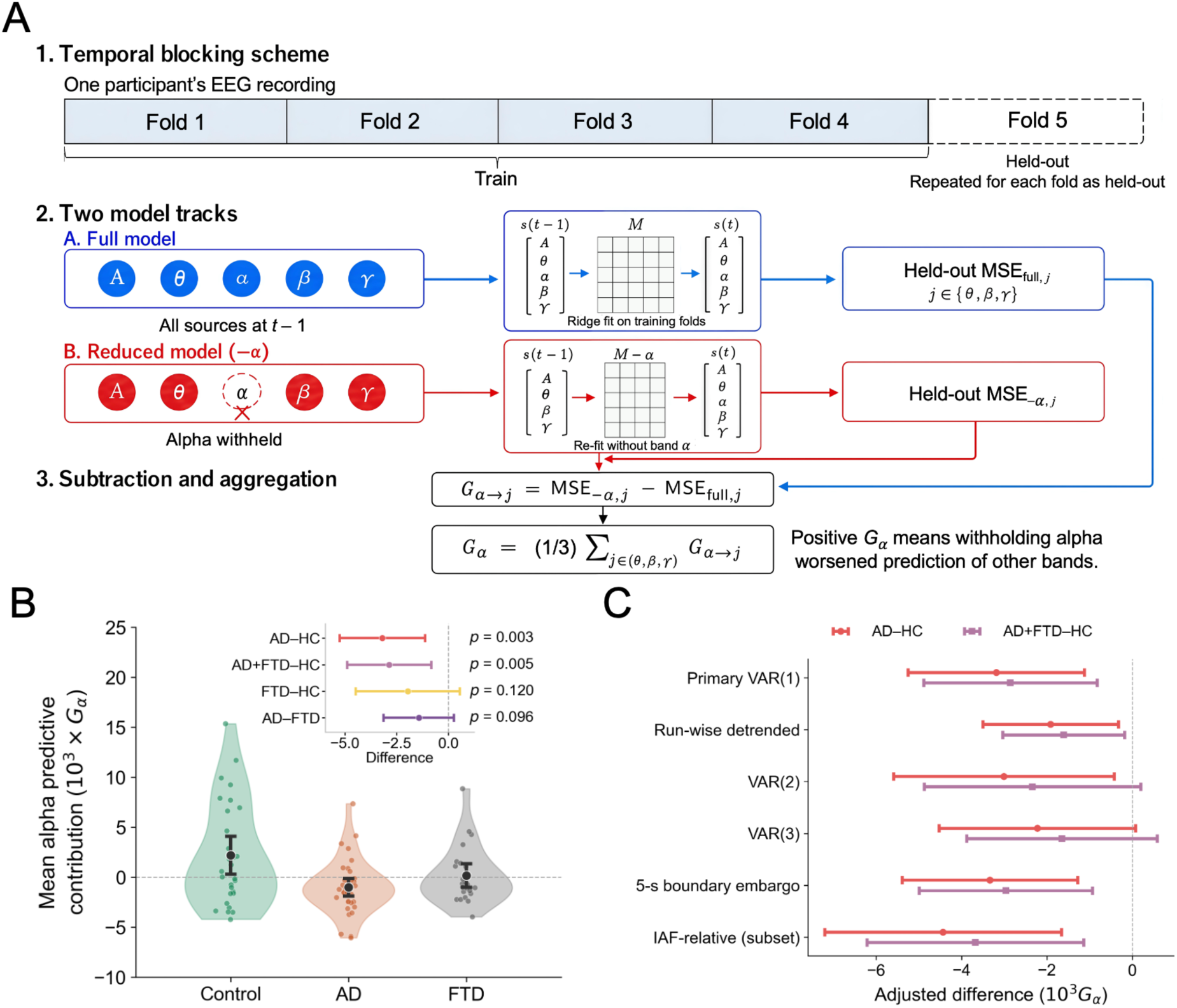
Alpha’s predictive contribution is reduced in AD. (A) Five-fold temporally blocked prediction. Each recording is divided into five contiguous folds. A ridge VAR(1) model, fitted to all pairs of consecutive windows in four folds, predicts the five states in each window of the held-out fold from all five states in the preceding window (A, aperiodic exponent); a reduced model refitted without lagged alpha predicts the same held-out observations. Each fold is held out in turn. *Gα* is the mean increase in held-out MSE for theta, beta and gamma when alpha is withheld. (B) Participant-level 10³*G_α_* with group means ± 1.96 standard errors (HC 2.191; AD −1.021; FTD 0.166). The inset shows age- and sex-adjusted contrasts with HC4 95% CIs: AD–HC −3.190 [−5.254, −1.126], *p* = 0.0035; AD+FTD–HC−2.857 [−4.892, −0.823], *p* = 0.0049; FTD–HC −1.967 [−4.474, 0.540], *p* = 0.12; AD–FTD −1.428 [−3.126, 0.270], *p* = 0.096. Negative values indicate that the alpha-withheld model predicted held-out data better. (C) AD–HC and pooled AD+FTD–HC differences for the primary model and five sensitivity analyses: run-wise detrending, VAR(2), VAR(3), a 5-s embargo, and IAF-relative bands (restricted to participants with a detected alpha peak: 48 AD and HC; 67 AD, FTD and HC). Bars show HC4 95% CIs.

Positive values of *G_α_* indicate that recent alpha activity improves out-of-sample prediction of the subsequent activity of theta, beta and gamma. Because the reduced model was refitted and evaluated on held-out data, *G_α_* measures incremental predictive information in the Granger sense [38, 39]. Values are reported as 10³*G_α_*. Five sensitivity analyses varied the specification: run-wise linear detrending, VAR(2), VAR(3), a 5-s embargo between training and test data, and bands defined relative to each participant’s individual alpha frequency (IAF) [20] (see details in Supplementary Methods 4–5).

### Adjustment for alpha expression and signal quality

To test whether the group difference in *G_α_* reflected alpha expression or signal quality, we fitted participant-level linear models with diagnosis, age and sex (model M0) and added alpha expression (M1), the two signal-quality measures (MQ) or both (M2). Retention was defined as the adjusted diagnosis coefficient as a percentage of the M0 coefficient, with 95% intervals from 5,000 diagnosis-stratified bootstrap samples. As a benchmark, the same models were applied to conditional alpha self-persistence (*P_α_*), that is, how strongly alpha in one window predicts alpha in the next window after accounting for the other four states, estimated as the alpha-to-alpha coefficient of the VAR(1) model fitted to the full recording. *P_α_* is expected to depend closely on alpha signal strength, because measurement noise in a weak alpha signal tends to reduce this coefficient. It therefore shows how an alpha-derived dynamic measure that depends on signal strength responds to the same adjustment. Attenuation was compared descriptively between the two measures. Model details are described in Supplementary Methods 6.

### Source, target and regional profiles

Three profiles tested whether the AD–HC difference was specific to alpha, to particular predicted rhythms or to posterior regions. Source profiles repeated the *G_α_* analysis with each rhythm (theta, alpha, beta or gamma) withheld in turn as the predictor (source) and averaged its contribution over the other three rhythms (targets). Target profiles separated alpha’s contribution to theta, beta and gamma, and regional profiles recomputed *G_α_* with alpha defined over anterior (F3, F4, Fz, C3, C4 and Cz) or posterior (O1, O2, P3, P4, Pz, T5 and T6) channels. Source and target profiles were analyzed with stacked regression models adjusted for age, sex and the M2 covariates, and regional profiles with region-specific expression and quality covariates, using participant-clustered standard errors [56] and omnibus tests of heterogeneity across sources, targets or regions; regional spatial weights were derived from controls. Alpha specificity was assessed by contrasting alpha with the mean of the other sources, with and without the M2 covariates (Supplementary Methods 7).

### Joint alpha phenotype and complementarity

We next asked whether alpha expression and *G_α_* provide overlapping or complementary information about AD. Alpha expression and *G_α_* were converted into deficit scores (*D*_E_ and *D*_G_) by standardizing each to the control mean and standard deviation and reversing the sign, so that positive values indicate lower expression or contribution; each control participant was scored against references that excluded that participant. AD–HC displacement in this two-dimensional space was tested with a joint Wald test adjusted for age and sex, with standard errors clustered by participant. Complementarity, that is, whether each measure added information for distinguishing AD from HC beyond the other, was tested with nested AD–HC logistic models compared by likelihood-ratio (LR) tests. Each deficit score was first added to a model containing age and sex; then, in two reciprocal tests, *D*_G_ was added to the model containing *D*_E_, and *D*_E_ to the model containing *D*_G_. The reciprocal tests address complementarity directly and were Holm-adjusted as a two-test family [57]; the first-step tests were secondary and are reported with nominal p values. Cohen’s d for each deficit score and for their equal-weight composite (the dual-alpha score; weights were not optimized), apparent areas under the receiver operating characteristic curve (AUC) and exploratory associations with MMSE were computed descriptively (Supplementary Methods 8).

### Exploratory circuit model

To examine whether expression and *G_α_* can change partly independently, we simulated a Jansen–Rit-family circuit [51–53] in which an alpha-generating column (α source in Figure 6A) provided delayed excitatory input to three downstream columns with different kinetics, each driven by independent stochastic input. We compared a reference condition with a 2% reduction in this column’s local recurrent excitation, a 50% reduction in its input to the downstream columns, and both changes together, using 24 paired 600-s realizations. The model settings were chosen in preliminary simulations and then fixed, and the reported results used a random seed not used in those simulations. Simulated signals were analyzed with the same spectral and predictive-contribution definitions as the EEG, applied to a single simulated signal rather than a spatial projection. Parameters, controls and alternative operating assumptions are given in Supplementary Methods 9 and Table S8.

### Statistical analysis

The primary outcome was the age- and sex-adjusted AD–HC difference in *G_α_*; pooled AD+FTD–HC, FTD–HC and AD–FTD contrasts were secondary. The sample size was determined by the public dataset, and no a priori power calculation was performed. Group differences were estimated with linear models. Ninety-five percent confidence intervals (CI) used HC4 heteroskedasticity-consistent standard errors [58], and p values were obtained from a restricted wild bootstrap with 10,000 Rademacher draws imposing the null hypothesis [59]. The primary test was not adjusted for multiplicity, and the two reciprocal complementarity tests were Holm-adjusted; other analyses were secondary and are interpreted from effect sizes and CIs, with nominal p values. Analyses were performed in Python 3.12 with NumPy, SciPy, pandas and statsmodels.

## Results

### Participant characteristics and alpha expression

Before testing alpha’s predictive contribution, we confirmed that this cohort showed the established reduction in alpha expression. Participant characteristics are summarized in Table 1. All 88 recordings yielded the five state trajectories, with a median of 802.5 retained 1-s windows per participant. Alpha expression was lower in both patient groups than in controls on conventional, PC1-weighted and periodic measures (Figure 2C, D; Table S1). For periodic alpha expression, the measure used in the subsequent analyses, the age- and sex-adjusted AD– HC difference was −0.975 dB (95% CI −1.579 to −0.371; *p* = 0.0033). AD and FTD did not differ significantly on any expression measure (Table S1).

**Table 1.** Participant characteristics

|  | HC (n = 29) | AD (n = 36) | FTD (n = 23) |
| --- | --- | --- | --- |
| Age, years, mean (SD) | 67.9 (5.4) | 66.4 (7.9) | 63.7 (8.2) |
| Women, n (%) | 11 (37.9) | 24 (66.7) | 9 (39.1) |
| MMSE, mean (SD) | 30.0 (0.0) | 17.8 (4.5) | 22.2 (2.6) |
| Retained 1-s windows, median [range] | 809 [738–976] | 814 [305–1274] | 695 [479–1006] |

AD, Alzheimer's disease; FTD, frontotemporal dementia; HC, healthy controls; MMSE, Mini-Mental State Examination; SD, standard deviation.

### Alpha’s predictive contribution was reduced in AD

We next addressed the primary question, whether alpha’s predictive contribution is reduced in AD. *G_α_* was positive on average in controls and negative on average in AD (mean 10³*G_α_*: HC 2.191, AD −1.021, FTD 0.166; Figure 3B). A negative *G_α_* means that the model refitted without alpha predicted held-out data better than the full model. This occurs when lagged alpha carries little or no information that generalizes to held-out data, so its fitted coefficients mainly add noise; it does not indicate reversed coupling. The adjusted AD–HC difference was −3.190 (95% CI −5.254 to −1.126; *p* = 0.0035), and the pooled AD+FTD–HC difference was also significant (Figure 3B). The FTD mean lay between the control and AD means, and the FTD–HC and AD–FTD differences were not significant, so these data do not establish whether the reduction is specific to AD. BIC favored a one-lag model in 86 of 88 participants (Additional file 1: Table S2), and the AD–HC difference remained negative in all five sensitivity analyses (Figure 3C). Estimates for the secondary contrasts and sensitivity analyses are given in Table S3.

### Adjustment for alpha expression and signal quality

Because weaker or noisier alpha could, in principle, lower *G_α_*, we then asked whether the reduction reflected alpha expression or signal quality. Adjustment for alpha expression retained 94.9% of the AD–HC difference in *G_α_*, adjustment for signal quality retained 67.3%, and joint adjustment retained 75.8% (M2: −2.419; 95% CI −4.633 to −0.205; *p* = 0.024; Figure 4A). The self-persistence benchmark also differed between AD and HC before adjustment (*p* = 0.0028), but the same three adjustments retained only 32.2%, 3.8% and 4.4% of this difference (M2: −3.283 in 10³*P_α_*; 95% CI −31.595 to 25.030; *p* = 0.81; Figure 4B). Bootstrap retention intervals, paired coefficient changes and further models, including additional adjustment for the overall correlation structure of the states, are reported in Tables S4–S5. In summary, most of the AD– HC difference in *G_α_* remained after adjustment for alpha expression and signal quality, whereas the same adjustment removed almost all the difference in alpha self-persistence.

**Figure 4.**
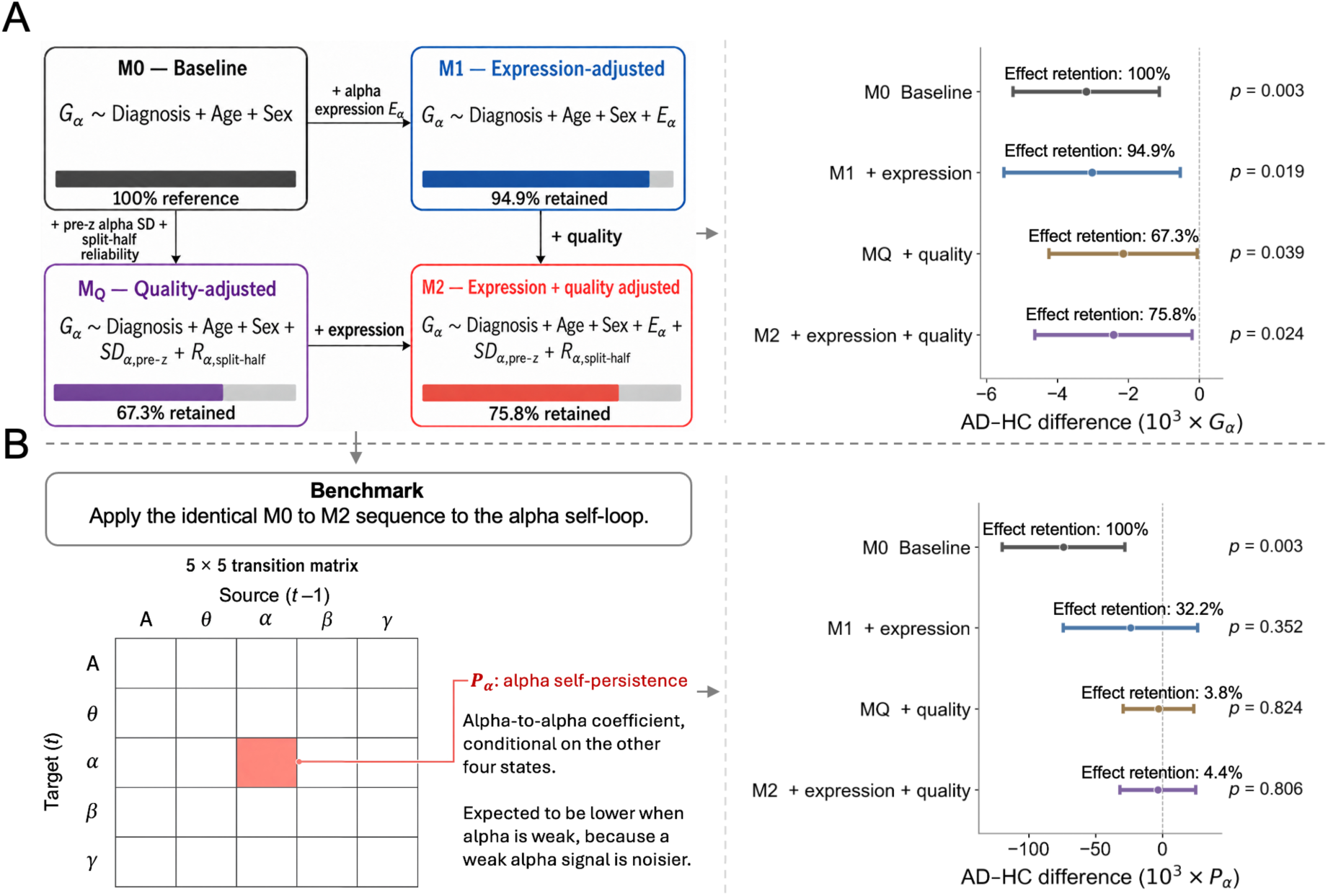
Adjustment for alpha expression and signal quality. (A) Left, the four adjustment models, with arrows marking the covariates added at each step. Formulas show the terms used to model *G_α_*, where *E_α_* is alpha expression (mean periodic alpha power above the aperiodic background), *SD_α_*, pre-z is alpha dynamic range (standard deviation of the alpha trajectory before z-scoring), and *R_α_*, split-half is the spatial split-half reliability of the alpha trajectory. Colored bars show the percentage of the M0 group difference retained after adjustment. Right, AD–HC difference in 10³*G_α_* adjusted for age and sex (M0) and additionally for alpha expression (M1), for alpha dynamic range and spatial split-half reliability (MQ), or for both (M2): −3.190, −3.026, −2.147 and −2.419, retaining 100%, 94.9%, 67.3% and 75.8% of M0 (bootstrap retention intervals for M1, MQ and M2: 44.0–131.2%, 16.9–98.5% and 18.6– 120.8%). (B) Benchmark with conditional alpha self-persistence (Pα). Left, the 5 × 5 VAR(1) transition matrix (A, aperiodic exponent), with source states in the preceding window (t − 1) as columns and target states in the current window (t) as rows. The highlighted alpha-to-alpha cell is *P_α_*, that is, how strongly alpha in one window predicts alpha in the next after accounting for the other four states. Because measurement noise in a weak alpha signal tends to reduce this coefficient, *P_α_* is expected to depend closely on alpha signal strength. It therefore shows how an alpha-derived dynamic measure with this dependence responds to the same adjustment as *G_α_*. Right, the same models applied to Pα: −73.863, −23.772, −2.783 and −3.283 in 10³ *P_α_* units, retaining 100%, 32.2%, 3.8% and 4.4% of M0. Unlike the Gα difference (A), the *P_α_* difference was almost entirely removed by adjustment.

### Source, target and regional profiles

To characterize the reduction further, we examined whether it was specific to alpha as a predictor, to particular predicted rhythms or to posterior regions. When each of the four rhythms was withheld in turn as the predictor (source), alpha showed the largest adjusted AD– HC reduction, and source effects differed nominally (omnibus F(3,64) = 2.836, *p* = 0.045; Figure 5A). The excess of alpha over the mean of the other sources was significant before expression and quality adjustment but not after it (Table S6). For each predicted (target) rhythm, the AD–HC difference in alpha’s contribution was negative, with no detectable difference among the three targets (*p* = 0.60; Figure 5B), and the reduction did not differ significantly between anterior and posterior alpha definitions (p = 0.34; Figure 5C).

**Figure 5.**
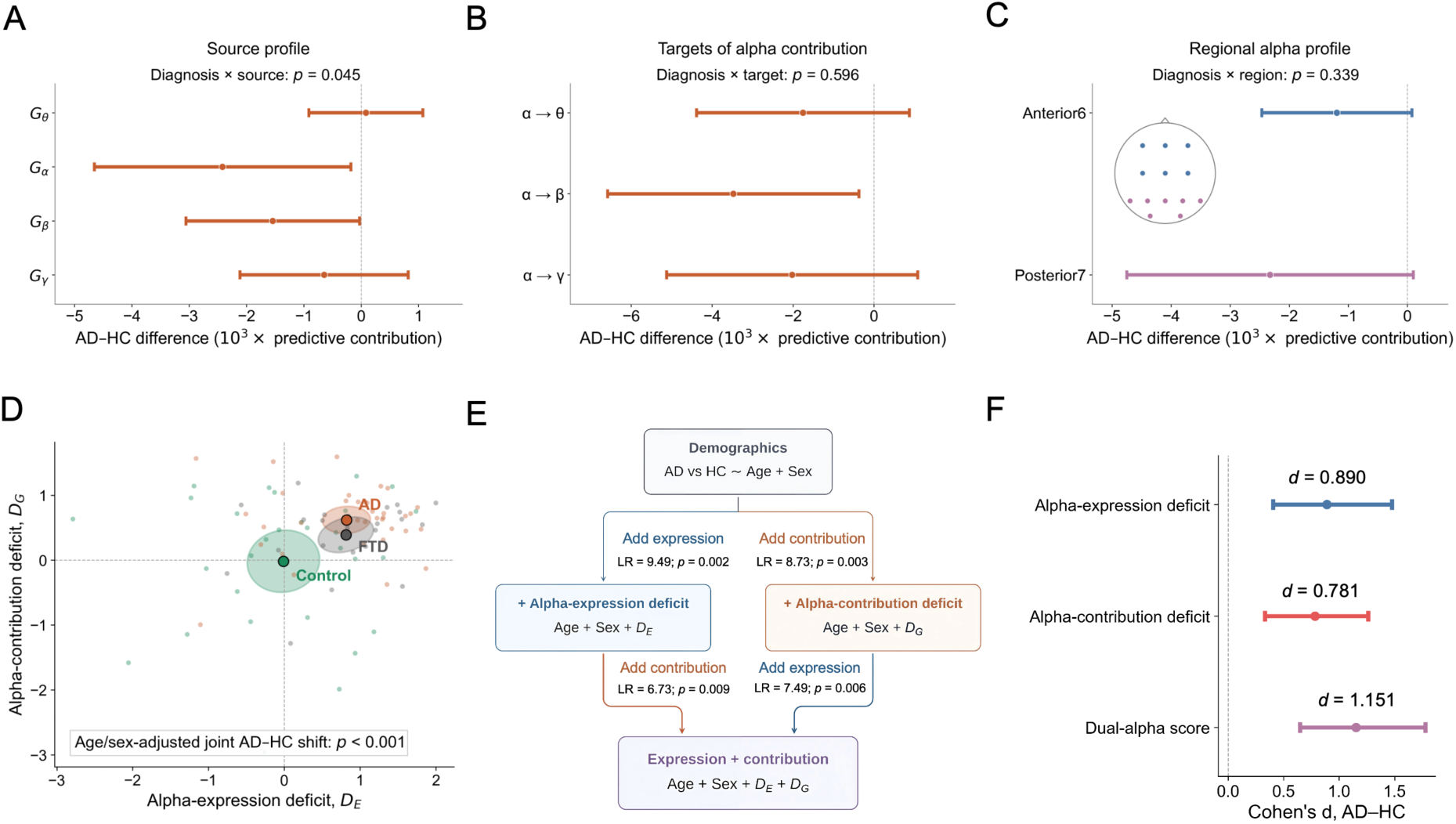
Source, target and regional profiles, and a joint alpha phenotype with complementary group information. (A–C) Points show AD–HC differences adjusted for age, sex, alpha expression and signal quality (M2; region-specific covariates in C), and bars show participant-clustered 95% CIs, on the 10³ scale. (A) Predictive contribution of each source rhythm averaged over the other three rhythms: theta 0.079, alpha −2.419, beta −1.544 and gamma −0.649 (omnibus F(3,64) = 2.836, *p* = 0.045; alpha minus the mean of the other sources, *p* = 0.14). (B) Alpha’s contribution to future theta, beta and gamma: −1.755, −3.477 and −2.024 (omnibus *p* = 0.60). (C) Alpha states defined over anterior (F3, F4, Fz, C3, C4 and Cz) and posterior (O1, O2, P3, P4, Pz, T5 and T6) channels: −1.195 and −2.326 (difference *p* = 0.34). (D) Expression deficit (*D*_E_) versus contribution deficit (*D*_G_), both referenced to controls, with positive values indicating lower expression or contribution. Points are participants, and large symbols are group means with approximate 95% bootstrap regions. AD and HC differed jointly (age- and sex-adjusted F(2,64) = 9.523, *p* < 0.001); FTD is shown for context. (E) Nested AD– HC logistic models. Adding expression or contribution to age and sex improved fit (LR = 9.49 and 8.73). Adding contribution to the expression model (LR = 6.73) and expression to the contribution model (LR = 7.49) both improved fit (Holm-adjusted *p* = 0.012 for each). Please note that *p* values shown in the panel are unadjusted. (F) Unadjusted Cohen’s d with 95% bootstrap intervals: expression deficit 0.890 [0.405, 1.476], contribution deficit 0.781 [0.331, 1.262] and their equal-weight combination 1.151 [0.648, 1.777].

### Alpha expression and *Gα* carried complementary group information

Since most of the *G_α_* reduction remained after adjustment for expression, we also asked whether the two measures carry complementary information about AD. When expression and *Gα* were considered together as deficit scores, AD and HC differed jointly (F(2,64) = 9.523, *p* < 0.001; Figure 5D). Adding either measure to age and sex improved the AD–HC logistic model, and in the reciprocal comparisons, adding *D*_G_ to the expression model (LR = 6.73) and adding *D*_E_ to the contribution model (LR = 7.49) both improved fit (Holm-adjusted *p* = 0.012 for each; Figure 5E). The expression deficit, the contribution deficit and their equal-weight combination had unadjusted Cohen’s d of 0.890, 0.781 and 1.151, respectively (Figure 5F). Apparent AUCs and exploratory associations with MMSE are reported in Table S7; the MMSE associations were not significant.

### Exploratory circuit model

Finally, we used the circuit model to ask whether expression and *G_α_* can change partly independently within a simple generative system. The reference circuit (Figure 6A) generated sustained alpha activity with window-to-window fluctuations (Figure 6B). Reducing local recurrent excitation in the alpha-generating column changed periodic alpha expression by −3.028 dB without decreasing *G_α_* (change in 10³*G_α_*, +6.158), whereas reducing the input from this column to the downstream columns lowered *G_α_* (−9.809) with a smaller change in expression (−0.474 dB). Applying both changes reduced expression and *G_α_* together (−3.329 dB and −10.697; Figure 6C). Paired bootstrap intervals are given in Table S8. These responses depended on the operating regime of the model, which was not calibrated to the human data.

**Figure 6.**
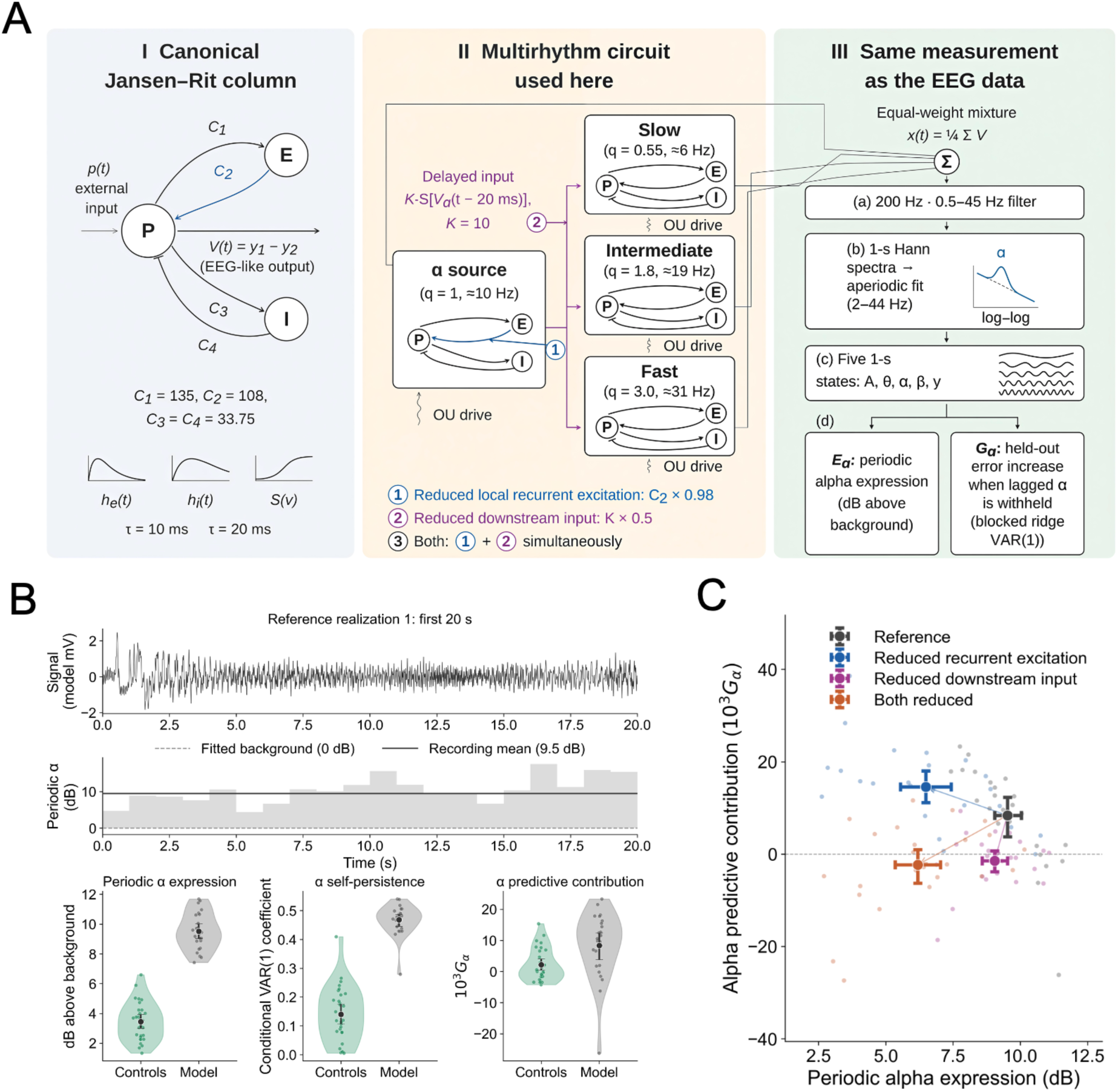
An exploratory circuit model shows that expression and *G_α_* can change partly independently. (A) Model structure. I, Jansen–Rit column with pyramidal (P), excitatory interneuron (E) and inhibitory interneuron (I) populations. II, an alpha-generating column (α source) sends delayed (20-ms) excitatory input to three downstream columns with different kinetics; every column receives independent stochastic drive. Interventions: ① reduced recurrent excitation in the alpha-generating column (*C*_2_ × 0.98); ② reduced input from this column to the downstream columns (K × 0.5); ③ both. III, the equal-weight voltage mixture is analyzed with the same spectral-state and blocked-ablation definitions as the EEG analysis. (B) First 20 s of one reference realization and its 1-s periodic alpha values, with a comparison of 29 controls and 24 model realizations (means and 95% bootstrap intervals). (C) Condition means across 24 paired realizations. Points show condition means; horizontal and vertical bars are marginal 95% percentile-bootstrap intervals across 24 realizations. Faint points show individual paired realizations and arrows indicate mean changes. Paired changes in expression (dB) and 10³*G_α_* were −3.028 and +6.158 for reduced recurrence, −0.474 and −9.809 for reduced downstream input, and −3.329 and −10.697 for both.

## Discussion

In this study, alpha’s predictive contribution (*G_α_*), which captures the temporal link from recent alpha fluctuations to the subsequent activity of theta, beta, and gamma, was lower in AD than in healthy controls (Figure 3B). This reduction was consistent in direction across the selected modeling choices (Figure 3C), and most of it remained after adjustment for alpha expression and signal quality (Figure 4A). By contrast, the same adjustment largely removed the group difference in conditional alpha self-persistence, that is, how strongly alpha carries over from one window to the next (Figure 4B). Alpha expression and *G_α_* each carried group information beyond the other (Figure 5E), and an exploratory circuit model showed that the two measures need not co-vary (Figure 6C). Together, these findings indicate that, at the group level, the alpha abnormality in AD has two partly nonredundant components: the well-established reduction in expression [1, 3] and a weaker temporal link between alpha and other rhythms. Most of the *G_α_* reduction in AD was not attributable to differences in alpha expression or signal quality. This needed to be tested because weaker expression could, in principle, lower *G_α_*. Expression was lower in AD on conventional, spatially weighted, and periodic measures (Figure 2C, D; Table S1), consistent with previous reports [1, 2, 5], and measurement error can bias a predictor’s estimated contribution [45, 46]. However, two observations argue against a purely expression-driven account. First, adjusting for expression alone retained 94.9% of the baseline *G_α_* difference, adjusting for two signal-quality measures (alpha dynamic range and spatial split-half reliability) retained 67.3%, and adjusting for both retained 75.8% (Figure 4A). By contrast, the same joint adjustment reduced the group difference in conditional alpha self-persistence, a benchmark quantity expected to depend closely on alpha signal strength, to 4.4% of its baseline value (Figure 4B). Second, reciprocal likelihood-ratio (LR) tests showed that expression and *G_α_* each added AD–HC group information beyond the other: adding *G_α_* to a logistic model containing expression improved fit, as did adding expression to a model containing *G_α_* (Figure 5E). *G_α_* therefore captures an aspect of the alpha abnormality in AD that expression measures do not.

Two other spectral features of AD, broadband aperiodic changes and a slower alpha peak, could in principle lower *Gα*, but neither is likely to explain the finding. Because the aperiodic exponent was modeled as a state, *G_α_* reflects alpha’s contribution beyond lagged broadband changes, which is important given that aperiodic findings in AD and other clinical conditions are inconsistent [5–8, 60]. In addition, because the alpha peak frequency is lower in AD [1], fixed band limits might assign part of the alpha activity to the theta band. Defining the bands around each participant’s own alpha peak, however, also yielded a reduced contribution in AD (Figure 3C). This check was possible only in participants with a clear alpha peak (20 of 36 with AD and 28 of 29 controls), but it suggests that the finding does not depend on fixed band limits. These findings complement previous work on interactions between rhythms. Cross-frequency studies in AD and MCI have reported altered phase–amplitude coupling and bispectral phase coupling, with inconsistent directions [26–29]. These measures index time-averaged or simultaneous association, whereas *G_α_* indexes lagged, directional dependence. Directed-connectivity studies do capture lagged dependence and have reported weaker or disturbed information flow in AD and MCI, but mainly between recording sites or sources within frequency bands [30, 40, 42, 43, 61]. The present results extend this directed perspective from interactions between locations to interactions between rhythms.

*G_α_* thus adds a dynamic dimension to the characterization of alpha in AD. Expression measures describe how strongly alpha is present on average, whereas *G_α_* describes how its moment-to-moment fluctuations relate to what other rhythms do next. Since *G_α_* is estimated out of sample, with the reduced model refitted and all other rhythms and the aperiodic background included, it quantifies the incremental information that recent alpha activity carries about the near future of the multi-rhythm system. Alpha activity has been linked to inhibitory gating of cortical processing and, together with beta, to feedback signaling, whereas gamma has been linked to feedforward signaling, mainly in visual cortex [21–25]. A reduced *G_α_* in AD is therefore consistent with a weakening of the coordinating role attributed to alpha. Among the four source rhythms, alpha showed the largest reduction, and its excess over the other sources was evident before adjustment for alpha expression and quality (Figure 5A; Table S6). Beta’s contribution was also modestly reduced, and the alpha reduction did not differ detectably across target rhythms or between anterior and posterior regions (Figure 5B, C). This pattern suggests a broader change in the temporal organization of multi-rhythm activity in AD, with alpha as its most affected source, consistent with altered microstate, meta-state, and hidden-state dynamics [35–37, 62, 63].

The circuit model complements these findings by showing that expression and predictive contribution can change partly independently within a simple generative system. In a Jansen– Rit-family circuit [51–53], reducing local recurrent excitation lowered alpha expression without decreasing *G_α_*, whereas reducing the input from the alpha-generating column to the downstream columns lowered *G_α_* with a smaller change in expression. Combining the two produced concurrent deficits in both measures, qualitatively resembling the pattern observed in AD (Figure 6C). Earlier neural mass models have related AD-associated excitation– inhibition changes to spectral abnormalities [64, 65]; our model instead asked whether the two alpha measures can dissociate. It was not fitted to patient data, and its behavior depended on operating assumptions (Table S8), as expected for simplified neural mass models [66]. It is therefore best viewed as a proof of principle rather than a mechanism of AD.

This study has some limitations. First, the analyses were based on a single public cohort with clinically defined groups recorded under a routine clinical EEG protocol [49]. Few openly available datasets include comparable diagnostic groups. One such resource, the BrainLat project, recorded 128-channel EEG across multiple sites with a different protocol and population [67], so applying the present analysis there would require harmonization across these differences. Assessing how *G_α_* generalizes across recording protocols, and in cohorts with biomarker confirmation and longitudinal follow-up, is an important next step. Such cohorts could also test whether *G_α_* tracks tau pathology, disease stage, and progression, as conventional alpha measures do [9, 11, 12]. Second, information on medication and vigilance, both of which influence resting alpha [17, 18, 20], was not available for analysis and should be incorporated in future recordings. Third, *G_α_* is a predictive measure: it shows that alpha fluctuations help predict later activity in other rhythms, but not which causal or anatomical pathway produces this link [38, 39]. Because all rhythmic states were derived from the same scalp recordings, they may also share signal components, and Granger-type measures can be biased when such components are measured with unequal signal-to-noise ratios or arise from a common input [68]. Our design limits these influences. *G_α_* was computed between rhythms rather than between recording sites, avoiding the apparent site-to-site links created by the instantaneous spread of activity across electrodes [69, 70]. Each rhythm was measured relative to its window’s own aperiodic background, removing broadband shifts shared across bands, and each prediction was conditioned on all other states. The AD–HC difference also largely persisted after adjustment for alpha signal quality (Figure 4A). These features reduce, but cannot fully exclude, contributions from shared components, so *Gα* is best interpreted as a predictive dependence among scalp-level rhythmic states. Future work could refine *G_α_* by comparing PCA with other ways of extracting rhythmic states, such as independent component analysis, generalized eigendecomposition, or source-space estimation [71]. Finally, the analyses focused on AD because the alpha expression deficit that motivated this study is best established in AD [1–3], whereas EEG findings in FTD are fewer and more heterogeneous [14, 16]. FTD therefore served as a clinical comparison group. With 23 participants, the FTD contrasts were less conclusive, and whether reduced *G_α_* also occurs in FTD or is specific to AD remains to be examined in larger samples.

## Conclusions

In resting-state EEG, the temporal link from recent alpha fluctuations to the subsequent activity of other rhythms, quantified as alpha’s predictive contribution, was weaker in AD than in healthy controls. Most of this reduction was not attributable to differences in alpha expression or signal quality, and expression and predictive contribution each carried group information beyond the other. At the group level, the alpha abnormality in AD is therefore better described by two partly nonredundant dimensions than by expression alone: how strongly alpha is expressed and how strongly its fluctuations are linked to subsequent multi-rhythm activity. An exploratory circuit model showed that these dimensions can change partly independently. Measuring the temporal link between alpha and other rhythms may complement conventional spectral markers in characterizing the neurophysiology of AD and warrants validation in independent, biomarker-defined cohorts.

## Supplementary Information

The online version contains detailed methods (spectral estimation and channel policy, aperiodic fitting and spatial states, expression and signal-quality measures, model specification and order assessment, source ablation and sensitivity analyses, covariate adjustment, profiles, complementarity and the exploratory circuit model; Supplementary Methods 1–9) and full numerical results (Tables S1–S8). The supplementary material is available at [].

## Declarations

### Ethics approval and consent to participate

This study is a secondary analysis of publicly available, de-identified data. Data collection for the original study was approved by the Scientific and Ethics Committee of AHEPA University Hospital, Aristotle University of Thessaloniki (protocol number 142/12-04-2023), and informed consent was obtained from all participants [49].

### Consent for publication

Not applicable.

### Availability of data and materials

The EEG data analyzed in this study are openly available in the OpenNeuro repository, https://doi.org/10.18112/openneuro.ds004504.v1.0.8 [50]. The analysis code will be available in the Open Science Framework at the time of publication.

## Funding

This work was supported in part by STI 2030-Major Projects (No. 2022ZD0208500), the Hong Kong Research Grant Council (No. GRF 12201421, No. CRF C4012-22G, SRFS2324-2S05) and Guangdong and Hong Kong Universities “1+1+1” Joint Research Collaboration Scheme (2025A0505000011), and Hong Kong Baptist University’s funding support (RC-SFCRG/23-24/SCI/06).

### Clinical trial number

Not applicable.

### Authors’ contributions

Jinwen Wei and Changsong Zhou conceived and designed the study; Jinwen Wei performed the analyses; Jinwen Wei and Changsong Zhou interpreted the results; Jinwen Wei drafted the manuscript. All authors read and approved the final manuscript.

## Data Availability

The EEG data analyzed in this study are openly available in the OpenNeuro repository, https://doi.org/10.18112/openneuro.ds004504.v1.0.8. The analysis code will be available in the Open Science Framework at the time of publication.

https://openneuro.org/datasets/ds004504/versions/1.0.8

## Acknowledgements

We thank the authors of OpenNeuro dataset ds004504 for making their recordings publicly available.

## List of abbreviations

AD: Alzheimer’s disease
AUC: area under the receiver operating characteristic curve
BIC: Bayesian information criterion
CI: confidence interval
*D*_E_: expression deficit score
*D*_G_: contribution deficit score
EEG: electroencephalography
FTD: frontotemporal dementia
*G_α_*: alpha’s predictive contribution
HC: healthy controls
HC4: heteroskedasticity-consistent covariance estimator, type 4
IAF: individual alpha frequency
LR: likelihood ratio
MCI: mild cognitive impairment
MMSE: Mini-Mental State Examination
MSE: mean squared error
NINCDS-ADRDA: National Institute of Neurological and Communicative Disorders and Stroke–Alzheimer’s Disease and Related Disorders Association
*P_α_*: conditional alpha self-persistence
PC1: first principal component
SD: standard deviation
VAR: vector autoregressive model

