## Supplementary methods and tables for "Beyond reduced alpha power: a weaker temporal link between alpha and other brain rhythms in Alzheimer’s disease"

This file gives the detailed methods summarized in the main text (Supplementary Methods 1–9) and the full numerical results (Tables S1–S8) for the cohort, measurement construction, temporal prediction, covariate adjustments, source and regional profiles, complementarity and exploratory circuit experiments. All secondary tests are exploratory unless explicitly stated otherwise.

Unless noted otherwise, effect intervals are HC4 Wald 95% confidence intervals and *p*-values are restricted wild-bootstrap values. Profile intervals are participant-clustered CR1 t-based intervals. Retention, correlation, effect-size and simulation intervals use percentile bootstrap resampling as specified for each analysis. The two reciprocal complementarity comparisons use Holm correction as a two-test family; other secondary *p*-values are nominal and exploratory. Predictive-contribution effects are displayed as 10³G; expression is displayed in dB where explicitly stated.

Supplementary Methods 1. Spectral estimation and high-frequency channel policy

Supplementary Methods 2. Aperiodic fitting, band values and spatial states

Supplementary Methods 3. Alpha-expression and signal-quality measures

Supplementary Methods 4. Transition model and model-order assessment

Supplementary Methods 5. Blocked alpha-source ablation, sensitivity analyses and inference

Supplementary Methods 6. Expression/quality adjustment and self-persistence benchmark

Supplementary Methods 7. Source, target and regional profiles

Supplementary Methods 8. Two-dimensional phenotype and reciprocal complementarity

Supplementary Methods 9. Exploratory circuit model

Table S1. Cohort and measurement definitions

Table S2. Transition order and adequacy

Table S3. Selected source-ablation sensitivities and scale context

Table S4. Expression/quality adjustment of Gα

Table S5. Conditional alpha self-persistence

Table S6. Source, target and regional profiles

Table S7. Complementarity and supporting analyses

Table S8. Exploratory circuit methods and results

### **Supplementary Methods**

These sections give the full technical details of the analyses summarized in the Methods of the main text. Throughout, i indexes participants, t one-second windows and c channels; f denotes frequency in Supplementary Methods 1–2 and the cross-validation fold in Supplementary Methods 5.

#### 1. Spectral estimation and high-frequency channel policy

Channels were aligned to Fp1, Fp2, F3, F4, C3, C4, P3, P4, O1, O2, F7, F8, T3, T4, T5, T6, Fz, Cz, and Pz. Fp1, Fp2, F7, and F8 were excluded from the primary rhythmic-state spatial basis because these anterior/peripheral electrodes were considered more susceptible to ocular and frontal artifact. This was a fixed channel-selection policy, not participant- or window-specific artifact rejection. Fp1/Fp2 remained available separately as high-frequency nuisance references. Theta, alpha, and beta used the remaining 15 channels. Gamma used the 11-channel subset F3, F4, C3, C4, P3, P4, O1, O2, Fz, Cz, and Pz: T3–T6 were excluded from its primary spatial compression. The aperiodic state retained the mean exponent across the 15-channel set. The band-specific gamma policy limits the direct contribution of temporal electrodes to the gamma state but does not establish that all retained signals are muscle-artifact-free. In particular, temporal-channel high-frequency activity can still affect the 2–44-Hz background fit, and hence the mean aperiodic exponent and lower-band residuals at those channels.

Signals were divided into non-overlapping one-second Hann windows, giving 1-Hz Fourier-bin spacing. Incomplete trailing windows and windows crossing an EEGLAB boundary event were not included. Original window indices and segment labels were preserved. No additional amplitude-based or aperiodic-fit-quality exclusion was applied; “retained windows” denotes temporal eligibility after the distributed preprocessing, not a newly performed comprehensive artifact screen. PSD bins from 1 through 44 Hz were retained, and the aperiodic fit used 2–44 Hz. Excluding the nominal 45-Hz cutoff bin does not imply an ideal, flat filter response through 44 Hz.

Before aperiodic fitting, high-frequency nuisance regression was applied separately at each 30–44-Hz bin. The nuisance reference $r_{it}\left( f \right)$ was the mean Fp1/Fp2 $\log_{10}$ power. A separate slope $b_{c}\left( f \right)$ was estimated for each target channel $c\in\{F3,F4,Fz\}$ from all pooled participant windows, without diagnosis labels. Corrected log power was

$$L_{itc}^{\mathrm{corr}}\left( f \right)=L_{itc}\left( f \right)-\hat{b}_{c}\left( f \right)\{r_{it}\left( f \right)-\overline{r}\left( f \right)\}.$$

Here $\overline{r}\left( f \right)$ is the pooled-window reference mean. The correction preserves the target’s pooled mean, not necessarily each participant’s mean. Participants with more windows contribute more observations to this nuisance fit; equal-participant weighting was used for the separate spatial covariance procedure below. The frontal reference was not assumed to measure temporal-muscle activity. T3–T6 were instead omitted from the primary gamma PC1. This is nuisance-variation removal, not identification of a pure EMG component; residual muscle activity and loss of genuine reference-correlated neural variation remain possible.

#### 2. Aperiodic fitting, band values and spatial states

For every window and retained channel, the aperiodic background was modelled as

$$\hat{L}_{\mathrm{ap}}^{\left( k \right)}\left( f \right)=b^{\left( k \right)}-\chi^{\left( k \right)}\log_{10}f.$$

The initial ordinary least-squares fit used all eligible 2–44-Hz bins. With residual $e^{\left( k \right)}\left( f \right)=L^{\mathrm{corr}}\left( f \right)-\hat{L}_{\mathrm{ap}}^{\left( k \right)}\left( f \right)$ and current eligibility weight $w^{\left( k \right)}\left( f \right)$, the mask threshold was

$$T^{\left( k \right)}=\left[ \frac{\sum_{f} w^{\left( k \right)}\left( f \right)e^{\left( k \right)}\left( f \right)^{2}}{\sum_{f} w^{\left( k \right)}\left( f \right)} \right]^{1/2}.$$

Bins with $e^{\left( k \right)}\left( f \right)>T^{\left( k \right)}$ were excluded from the next fit. Two mask updates yielded three fits. All frequencies were reevaluated at each update, allowing re-entry rather than imposing cumulative exclusion. If an update left fewer than 10 eligible bins, all fitting bins were restored for that update. This was a fitting fallback, not rejection of the window. Only positive residuals were masked to limit upward displacement of the background by spectral peaks. The procedure estimated offset and exponent only, without Gaussian peak fitting, and was not a FOOOF/specparam implementation.

The offset and exponent of the final fit defined that background. The final residual $R_{itc}\left( f \right)=L_{itc}^{\mathrm{corr}}\left( f \right)-\hat{L}_{ap,itc}\left( f \right)$ was calculated at all retained frequencies, including those excluded during background fitting. Multiplication by 10 gives the dB ratio of observed power to fitted background power. Residual means were taken over upper-exclusive bands [4,8), [8,13), [13,30), and [30,45) Hz, corresponding to the observed 4–7, 8–12, 13–29, and 30–44-Hz bins. Delta [2,4) Hz was retained diagnostically but omitted from the five-state temporal model because it comprises only two bins at the lower fitting boundary. This limits the measurement scope and does not imply that delta slowing is unimportant.

For each rhythmic band, a participant-specific channel mean and covariance were first calculated from eligible window-level maps. These means and covariances were averaged with one equal weight per participant. The leading eigenvector of the average covariance defined a fixed, unit-norm PC1 loading, with sign oriented toward the positive all-channel direction. Covariance dimension was 15 × 15 for theta, alpha, and beta and 11 × 11 for gamma. Centered maps were projected onto these loadings. The five trajectories were

$$\mathbf{s}_{i}\left( t \right)=\left[ A_{i}\left( t \right),\theta_{i}\left( t \right),\alpha_{i}\left( t \right),\beta_{i}\left( t \right),\gamma_{i}\left( t \right) \right]^{\mathsf{T}},$$

where $A_{i}\left( t \right)$ is the mean aperiodic exponent. Each trajectory was standardized over the participant’s full recording. Spatial loadings, nuisance regression, and temporal standardization were not re-estimated within cross-validation folds; only the predictive-model coefficients and predictor/target centering were estimated within training folds.

#### 3. Alpha-expression and signal-quality measures

The primary static expression measure, $E_{global,periodic}$, was the mean alpha residual across all windows and all 15 channels, before temporal standardization. It was stored in $\log_{10}$ units and displayed in dB above background after multiplication by 10. Posterior periodic expression used O1, O2, P3, P4, and Pz. Conventional posterior absolute alpha used spectra before nuisance correction and aperiodic subtraction: linear power was summed over 8–12 Hz, averaged across windows and those five channels, and then log-transformed. Posterior relative alpha was the average alpha-to-1–44-Hz linear-power ratio and was retained as a supplementary alternative.

The illustrative alpha-PC1 expression score was the recording mean of the centered, pre-standardization alpha PC1 trajectory, multiplied by 10. Its dB-scaled PC1 units are distinct from both conventional absolute power and the channel-mean expression used in the adjustment and complementarity analyses. These measures and their diagnostic contrasts were reported separately.

Alpha dynamic range $V_{i}$ was the standard deviation of the pre-standardization alpha trajectory. Spatial split-half reliability was computed over 100 fixed balanced 7/8-channel partitions of the 15-channel alpha maps. For each partition, the two channel-mean trajectories were correlated; correlations were averaged on the Fisher-z scale and transformed back. The Fisher-z-transformed summary, $R_{i}$, entered the adjustment models. Dynamic range and split-half agreement may contain biological as well as measurement variation and were not treated as pure measurement-error variables.

#### 4. Transition model and model-order assessment

The primary temporal specification was ridge VAR(1), with a one-second lag and the coefficient matrix oriented as future target by lagged source. A lagged observation was eligible only if the target and every required preceding window were consecutive in original time within the same recording segment. No lag crossed a rejected boundary window, temporal gap, or segment boundary. Centered training matrices $X$ and $Y$ gave

$$\hat{B}=\left( X^{\mathsf{T}}X+0.10nI \right)^{-1}X^{\mathsf{T}}Y, M=\hat{B}^{\mathsf{T}},$$

where $n$ is the number of training observations. Thus, the ridge penalty was fixed on the mean-squared-error scale rather than as a constant summed-error matrix penalty. This normalizes shrinkage for training size but does not remove all finite-sample effects of recording length.

VAR(1), VAR(2), and VAR(3) were compared on exactly the target windows eligible for VAR(3), so target times and values were identical across candidate orders. Approximate Gaussian AIC/BIC used the residual covariance and nominal transition-parameter count $25p$. Because estimates were ridge-regularized, this was a working information-criterion diagnostic, not an exact effective-degrees-of-freedom likelihood comparison. It assessed a common specification rather than selecting an individual model order for each participant. Full-record VAR(1) matrices were saved separately for the self-persistence benchmark and were not reused for held-out prediction. Spectral radii and ten-lag Ljung–Box diagnostics, including run-wise detrended fits, were retained as supplementary adequacy checks.

#### 5. Blocked alpha-source ablation, sensitivity analyses and inference

Valid lagged observations were ordered by original time and divided into five contiguous test folds. For each fold, the full ridge model used all five lagged states and the reduced model was independently refitted after removing lagged alpha. Both models used training-fold centering and predicted identical held-out observations. Alpha coefficients were not simply set to zero after fitting the full model.

For participant i, fold f, and future target j,

$$G_{\alpha\to j,i}=\frac{1}{F}\sum_{f=1}^{F} \left( \mathrm{MSE}_{-\alpha,i,f,j}-\mathrm{MSE}_{full,i,f,j} \right),$$

$$G_{\alpha,i}=\frac{G_{\alpha\to\theta,i}+G_{\alpha\to\beta,i}+G_{\alpha\to\gamma,i}}{3}.$$

The alpha target equation was excluded from the average; the aperiodic predictor remained in both models. Positive $G_{\alpha}$ indicates that withholding alpha worsens held-out prediction. Negative values mean that the refitted reduced model predicts better, not that physiological coupling is inhibitory or reversed. $G_{\alpha}$ is a difference in standardized squared-state prediction error, not a percentage or anatomical information-flow estimate. Results are displayed as ${10}^{3}G_{\alpha}$ for readability.

Participant-level linear models included age and sex. The primary AD–HC comparison was followed by pooled AD+FTD–HC, FTD–HC, and AD–FTD contrasts. Predictor scaling was performed within the relevant analysis sample. Confidence intervals were HC4 Wald intervals, $\hat{\beta}\pm1.96\mathrm{SE}_{HC4}$. Diagnosis p-values came from a two-sided restricted Rademacher wild bootstrap of the unstudentized coefficient, imposing the diagnosis null, with 10,000 draws, base seed 20260727, and deterministic analysis-specific streams. The plus-one estimate $\left( 1+\text{exceedances} \right)/\left( 10000+1 \right)$ prevented zero p-values. Values below 0.001 were reported as $p<0.001$. HC4 intervals and bootstrap p-values use different procedures and can disagree near the null. The primary baseline fit was reused in the adjustment analysis after checking matching samples, outcomes, covariates, and inference settings.

Selected sensitivity analyses are reported alongside the primary VAR(1) reference in Figure 3C of the main text and in Tables S3a–S3b. The six displayed specifications are primary VAR(1), run-wise linear detrending, VAR(2), VAR(3), a five-second fold-boundary embargo, and IAF-relative bands in participants with a qualifying alpha peak. The five sensitivity specifications address slow trends, temporal history, train/test proximity, and band definition. Participant endpoints were re-estimated using the same blocked full-versus-reduced model comparison. In higher-order models, lagged alpha was removed at all included lags. The five-second embargo removed training lag pairs that shared observations with the held-out fold and additionally excluded training target windows within five seconds of a held-out target window in the same recording segment. These are selected comparisons, not an exhaustive inventory of all analyses generated by the computational workflow.

IAF was the most prominent qualifying peak of the median posterior periodic spectrum from O1, O2, P3, P4, Pz, T5, and T6 within 6–14 Hz, requiring prominence ≥0.05 $\log_{10}$ units. No peak was imputed. Relative theta covered 4 Hz to IAF−2 Hz, alpha IAF±2 Hz, and beta IAF+2 to 30 Hz; gamma retained the fixed band and 11-channel basis. The IAF-relative analysis was restricted to participants with a detected peak, with identical eligible participants used for the full and alpha-withheld models. Because its sample differs from the full-cohort primary analysis, differences between these estimates cannot be attributed solely to changing the band definition.

For scale context, the full model’s mean held-out MSE improvement over a training-fold mean-only predictor was averaged across theta, beta, and gamma. Alpha’s signed relative share was $G_{\alpha}$ divided by that improvement, defined only for denominators above ${10}^{-12}$. Small denominators can make this ratio unstable, and it is not restricted to [0,1]. It was summarized descriptively alongside eligibility counts and lag support, not promoted to a second primary endpoint.

Eligibility for this descriptive ratio required only a positive mean held-out improvement over the training-mean predictor (apart from the numerical tolerance); it did not require a statistically significant or clinically meaningful predictive improvement.

#### 6. Expression/quality adjustment and self-persistence benchmark

The participant-level $G_{\alpha}$ endpoint was held fixed while between-participant covariates were varied. With diagnosis $D_{i}$, age, and sex, the models were

$$\begin{matrix} M0 & :G_{\alpha,i}=\beta_{0}+\beta_{D}D_{i}+\beta_{A}\mathrm{Age}_{i}+\beta_{S}\mathrm{Sex}_{i}+\varepsilon_{i}, \\ M1 & :M0+\beta_{E}E_{i}, \\ MQ & :M0+\beta_{V}V_{i}+\beta_{R}R_{i}, \\ M2 & :M0+\beta_{E}E_{i}+\beta_{V}V_{i}+\beta_{R}R_{i}. \end{matrix}$$

Here $E_{i}=E_{global,periodic,i}$. M1 and MQ were parallel branches from M0, not a stepwise sequence requiring monotonic attenuation. Posterior periodic and relative expression were supplementary alternatives. M2b additionally included log state-correlation condition number; M3 added diagnosis × expression to M2 as an interaction diagnostic.

The same sequence was applied to conditional alpha self-persistence, $P_{\alpha,i}=M_{\alpha,\alpha,i}$, from the full-record VAR(1). This in-sample conditional coefficient is not a univariate autocorrelation or a direct measure of alpha power. It was a comparative attenuation benchmark, not a negative control matched to the out-of-sample properties of $G_{\alpha}$.

Retention was $100\hat{\beta}_{D,m}/\hat{\beta}_{D,M0}$. Its uncertainty and that of the coefficient change were calculated from 5,000 diagnosis-stratified participant bootstrap samples, fitting baseline and adjusted models on the same sampled participants. Ratio intervals were withheld when the baseline bootstrap interval crossed zero or fewer than 95% of draws preserved its observed sign; coefficient-change intervals were retained. Retention is a descriptive coefficient ratio, not a percentage of a biological mechanism. No formal between-endpoint test of attenuation was performed. AD–HC was the principal comparison; other contrasts and model variants are supplementary.

#### 7. Source, target and regional profiles

Predictive contribution was calculated for each withheld source $s\in\{\theta,\alpha,\beta,\gamma\}$ by averaging its reduced-minus-full held-out error over the other three rhythmic targets. Four source outcomes per participant were stacked in a block-diagonal regression with a source-specific intercept, diagnosis effect, and covariate slopes. Participant-clustered CR1 covariance retained their within-person dependence; coefficient intervals used the cluster-based t distribution. The three-degree-of-freedom diagnosis-by-source test evaluated equality of the four diagnosis coefficients. The principal profile used the same alpha-focused M2 covariates for every source, asking about residual differences after adjustment for measured alpha expression/quality, not adjustment for every source’s own signal quality. The age/sex-only M0 profile and alpha-minus-mean-other-sources contrast were supplementary.

The three alpha-to-target outcomes were analysed analogously, with a two-degree-of-freedom diagnosis-by-target test and a direct beta-minus-gamma contrast. Regional analyses changed only the alpha state: anterior6 used F3, F4, Fz, C3, C4, and Cz; posterior7 used O1, O2, P3, P4, Pz, T5, and T6. Regional PC1 loadings used equal-participant covariance within the 29 HC participants, unlike the all-participant global bases. This HC-reference choice was not selected to maximize group separation, but it is diagnosis-informed reference construction. Other states remained global, including the revised gamma state. Regional trajectories were standardized and source ablation rerun. Region-specific expression and quality covariates entered the paired model, and the regional comparison tested equality of the anterior and posterior diagnosis effects. Non-significant contrasts were not interpreted as equivalence.

#### 8. Two-dimensional phenotype and reciprocal complementarity

Expression and contribution were expressed as HC-referenced deficit scores,

$$D_{E,i}=-\frac{E_{i}-\mu_{E,HC}}{\sigma_{E,HC}}, D_{G,i}=-\frac{G_{\alpha,i}-\mu_{G,HC}}{\sigma_{G,HC}}.$$

Larger values indicate lower expression or contribution. For each HC participant, component reference means and SDs excluded that participant; AD and FTD used the full HC reference. An equal-weight composite, $0.5D_{E}+0.5D_{G}$, was subsequently scaled using the fixed full-HC composite reference derived without leave-one-out scoring. Consequently, the final scored HC components and composite need not have exactly zero mean and unit SD. The weights were not optimized from diagnosis labels.

For AD and HC, a joint stacked regression of $D_{E}$ and $D_{G}$ assigned separate intercepts, diagnosis effects, and age/sex slopes to the two outcomes. A participant-clustered CR1 Wald F-test evaluated the joint null that both diagnosis coefficients were zero. This tests displacement, not complementarity. Group centroids were unadjusted; approximate 95% bivariate centroid regions used the covariance of 5,000 within-group bootstrap means, holding the score reference fixed. FTD provided descriptive context only.

Complementarity was tested separately through nested AD–HC logistic models: age+sex; age+sex+D_E; age+sex+D_G; and age+sex+D_E+D_G. Each one-variable addition was evaluated by a one-degree-of-freedom likelihood-ratio test. The two reciprocal comparisons of the joint model with each single-component model tested incremental group information, not independence of the underlying biological processes, and were treated as a two-test family with Holm adjustment. Logistic coefficient summaries used HC3 covariance. Cohen’s d and 95% group-stratified bootstrap intervals summarized unadjusted group differences for the components and composite. Supporting analyses included descriptive component correlations, first-versus-second-half ICC(2,1), AUC and paired AUC differences, and MMSE regressions. These exploratory analyses did not validate an individual-level diagnostic score.

#### 9. Exploratory circuit model

A Jansen–Rit-family circuit (main-text references [51–53]) was used to examine whether alpha expression and $G_{\alpha}$ must change together. As shown in main-text Figure 6A, the model has three parts: a canonical Jansen–Rit column (I), four such columns connected into a multirhythm circuit (II), and the same measurement pipeline as for the EEG (III). In this section, $t$ denotes continuous time and $k$ indexes columns. Parameter values are also listed in Table S8a.

*Canonical column (Figure 6A, I).* Each column contains pyramidal (P), excitatory-interneuron (E) and inhibitory-interneuron (I) populations. Incoming firing is converted into postsynaptic potentials by excitatory and inhibitory kernels,

$$h_{e}\left( t \right)=H_{e} a t e^{-at}, h_{i}\left( t \right)=H_{i} b t e^{-bt}, t\geq0,$$

and mean membrane potential is converted into firing rate by the sigmoid

$$S\left( v \right)=\frac{2e_{0}}{1+exp\left[ r\left( v_{0}-v \right) \right]}.$$

Writing $y_{0}$ for the postsynaptic potential that pyramidal firing evokes in both interneuron populations, and $y_{1}$ and $y_{2}$ for the excitatory and inhibitory postsynaptic potentials on pyramidal cells, each column obeys

$$\begin{matrix} \ddot{y}_{0} & =H_{e}a S\left( y_{1}-y_{2} \right)-2a\dot{y}_{0}-a^{2}y_{0}, \\ \ddot{y}_{1} & =H_{e}a\left[ p\left( t \right)+C_{2}S\left( C_{1}y_{0} \right) \right]-2a\dot{y}_{1}-a^{2}y_{1}, \\ \ddot{y}_{2} & =H_{i}b C_{4}S\left( C_{3}y_{0} \right)-2b\dot{y}_{2}-b^{2}y_{2}, \end{matrix}$$

with EEG-like output $V\left( t \right)=y_{1}\left( t \right)-y_{2}\left( t \right)$. Here $p\left( t \right)$ is the external input to the pyramidal population, $C_{1}$ and $C_{2}$ scale the excitatory feedback loop (pyramidal cells to excitatory interneurons and back), and $C_{3}$ and $C_{4}$ scale the inhibitory loop. Reference values were $H_{e}=3.25$ mV, $H_{i}=22$ mV, $a=100$ s⁻¹ and $b=50$ s⁻¹ (time constants $1/a=10$ ms and $1/b=20$ ms), $C_{1}=135$, $C_{2}=108$, $C_{3}=C_{4}=33.75$, $2e_{0}=5$ s⁻¹, $v_{0}=6$ mV and $r=0.56$ mV⁻¹.

*Multirhythm circuit (Figure 6A, II).* The circuit comprised an alpha-generating column, called the source column below (not to be confused with the source rhythms of Supplementary Methods 7), and three downstream columns, labeled slow, intermediate and fast. Each column $k$ had a kinetic factor $q_{k}$ that multiplied both rate constants and both gains,

$$a_{k}=q_{k}a, b_{k}=q_{k}b, H_{e,k}=q_{k}H_{e}, H_{i,k}=q_{k}H_{i},$$

with $q_{k}=1$, 0.55, 1.8 and 3.0 for the source, slow, intermediate and fast columns. Because $H_{e}/a$ and $H_{i}/b$ are unchanged, the kernels are only rescaled in time, $h_{e,k}\left( t \right)=q_{k}h_{e}\left( q_{k}t \right)$, so the columns differ in speed but not in synaptic gain. The approximate frequencies in Figure 6A (about 10, 6, 19 and 31 Hz) are nominal labels rather than measured peaks. Each column received its own Ornstein–Uhlenbeck (OU) drive,

$$d\xi_{k}=-\frac{\xi_{k}-\mu_{k}}{\tau_{\xi}} dt+\sigma_{\xi}\sqrt{\frac{2}{\tau_{\xi}}} dW_{k},$$

with correlation time $\tau_{\xi}=0.5$ s, stationary standard deviation $\sigma_{\xi}=22$ s⁻¹, mean $\mu_{k}=100$ s⁻¹ for the source and 90 s⁻¹ for the downstream columns, and independent Wiener processes $W_{k}$. The source received only its own drive, whereas each downstream column also received delayed excitatory input proportional to the source’s firing rate:

$$p_{\alpha}\left( t \right)=\xi_{\alpha}\left( t \right), p_{k}\left( t \right)=\xi_{k}\left( t \right)+K S\left[ V_{\alpha}\left( t-\delta\right) \right],$$

with $K=10$ and $\delta=20$ ms. Downstream columns did not project back to the source.

*Measurement (Figure 6A, III).* The simulated signal was the equal-weight mixture of the four column outputs,

$$x\left( t \right)=\frac{1}{4}\left[ V_{\alpha}\left( t \right)+V_{\text{slow}}\left( t \right)+V_{\text{int}}\left( t \right)+V_{\text{fast}}\left( t \right) \right],$$

which was processed as the EEG: (a) resampled from 1 kHz to 200 Hz and band-pass filtered at 0.5–45 Hz with a fourth-order zero-phase Butterworth filter; (b) transformed into 1-s Hann-window spectra, with the aperiodic background fitted over 2–44 Hz in log–log coordinates using the same iterative masking as for the EEG (Supplementary Methods 2); (c) summarized in each window as five states, the aperiodic exponent (A) and periodic theta (4–7 Hz), alpha (8–12 Hz), beta (13–29 Hz) and gamma (30–44 Hz) activity, each standardized over the recording; and (d) reduced to the two alpha measures. Alpha expression $E_{\alpha}$ was the recording-mean periodic alpha power in dB above the background, and $G_{\alpha}$ was the increase in held-out MSE for theta, beta and gamma when lagged alpha was withheld, using the same five-fold blocked ridge VAR(1) ablation as for the EEG (Supplementary Methods 5). Conditional alpha self-persistence $P_{\alpha}$ was the alpha-to-alpha coefficient of the ridge VAR(1) fitted to the full recording. Because the model produces a single signal, no spatial PC1 projection was applied.

*Interventions and simulation.* Four paired conditions were compared (Figure 6A, ①–③): the reference; reduced local recurrent excitation in the source column (① $C_{2}\times0.98$, that is, 108 → 105.84, with downstream columns unchanged); reduced downstream input (② $K\times0.5$, that is, 10 → 5); and both changes together (③). Lowering $K$ reduces both the mean and the fluctuating parts of the source-derived input; a control that restores the mean input is described below. Within each paired realization, all conditions shared the same OU drives and initial states, with all column variables starting at zero. The OU drives were generated with their exact autoregressive discretization at a 1-ms step, starting from the stationary distribution. The column equations were integrated with a fourth-order Runge–Kutta scheme with the same 1-ms step, holding the drive constant within each step, and the first 20 s were discarded as burn-in, leaving 600 s per realization. The configuration was selected in preliminary simulations, informed by their outcomes, and was then fixed and evaluated in 24 paired realizations with seed 2026092301, which had not been used in those preliminary simulations.

*Controls and operating assumptions.* Four controls each changed one operation relative to the reference (Table S8a): removing the downstream input ($K=0$); halving $K$ while restoring the mean downstream input; reducing the source’s weight in the observed mixture from 0.25 to 0.175 without changing the circuit; and circularly shifting the source firing-rate sequence by 30 s before it reached the downstream columns. A 5 × 5 grid crossed $C_{2}$ multipliers of 0.96–1.00 (steps of 0.01) with $K$ multipliers of 0–1 (steps of 0.25), with four paired 300-s realizations per cell. Eleven operating-assumption settings were each evaluated with eight paired 400-s realizations: OU correlation times of 150, 250 and 750 ms; delays of 0 and 40 ms; a 0.5-ms integration step; equal kinetics in all columns; source mixture weights of 1, 0.5 and 0.125, with the remainder shared equally by the downstream columns; and a repeated baseline. Results for these settings, together with source spectra, are given in Table S8.

*Comparison with controls and uncertainty.* Reference-model values were compared descriptively with those of the 29 controls. Because the model has a single mixture signal rather than scalp channels, the human values were channel-mean periodic expression and the $P_{\alpha}$ and $G_{\alpha}$ obtained from spatial-PC1 states, whereas the model values came from the mixture. This comparison checks that the model operates on a comparable measurement scale; it is not a calibration to the controls. Condition means and paired changes were summarized with 5,000 percentile-bootstrap resamples of realizations, and human and model means in the measurement check were resampled separately. These intervals describe simulation and sampling variability under the chosen model, not uncertainty about a disease mechanism. The simulations were exploratory, depended on the chosen operating regime, and were not fitted to patients.

### Table S1. Cohort and measurement definitions

#### S1a. Participant and recording characteristics

Table S1a. Participant characteristics and available recording duration.

| Group | n | Age, mean (SD) | Women, n (%) | MMSE, mean (SD) | Derivative seconds, median [range] | Retained 1-s windows, median [range] |
| --- | --- | --- | --- | --- | --- | --- |
| HC | 29 | 67.9 (5.4) | 11 (37.9%) | 30.0 (0.0) | 814.60 [748.48, 989.88] | 809 [738, 976] |
| AD | 36 | 66.4 (7.9) | 24 (66.7%) | 17.8 (4.5) | 820.03 [306.10, 1281.72] | 814 [305, 1274] |
| FTD | 23 | 63.7 (8.2) | 9 (39.1%) | 22.2 (2.6) | 699.14 [479.10, 1014.50] | 695 [479, 1006] |

Derivatives need not equal the original acquisition duration. Retained duration equals the number of one-second windows. No new artifact-quality exclusion is implied.

#### S1b. Alpha-expression measures and adjusted contrasts

##### Posterior absolute alpha (dB)

Table S1b(i). Conventional posterior absolute-alpha contrasts.

| Contrast | n | Adjusted difference [95% HC4 CI] | Wild *p* |
| --- | --- | --- | --- |
| AD–HC | 65 | −3.103 [−4.439, −1.767] | <0.001 |
| AD+FTD–HC | 88 | −3.067 [−4.309, −1.824] | <0.001 |
| FTD–HC | 52 | −3.127 [−4.785, −1.469] | <0.001 |
| AD–FTD | 59 | −0.462 [−1.930, 1.005] | 0.540 |

##### Alpha-PC1 expression (dB-scaled PC1 units)

Table S1b(ii). Centered alpha-PC1 expression contrasts.

| Contrast | n | Adjusted difference [95% HC4 CI] | Wild *p* |
| --- | --- | --- | --- |
| AD–HC | 65 | −4.046 [−6.469, −1.622] | 0.002 |
| AD+FTD–HC | 88 | −4.141 [−6.342, −1.941] | <0.001 |
| FTD–HC | 52 | −4.209 [−6.784, −1.634] | 0.002 |
| AD–FTD | 59 | −0.559 [−2.767, 1.648] | 0.610 |

##### Global periodic alpha (dB)

Table S1b(iii). Global channel-mean periodic-alpha contrasts.

| Contrast | n | Adjusted difference [95% HC4 CI] | Wild *p* |
| --- | --- | --- | --- |
| AD–HC | 65 | −0.975 [−1.579, −0.371] | 0.003 |
| AD+FTD–HC | 88 | −1.008 [−1.557, −0.459] | <0.001 |
| FTD–HC | 52 | −1.037 [−1.675, −0.400] | 0.002 |
| AD–FTD | 59 | −0.106 [−0.647, 0.436] | 0.701 |

##### Posterior periodic alpha (dB)

Table S1b(iv). Posterior periodic-alpha contrasts.

| Contrast | n | Adjusted difference [95% HC4 CI] | Wild *p* |
| --- | --- | --- | --- |
| AD–HC | 65 | −1.515 [−2.328, −0.701] | <0.001 |
| AD+FTD–HC | 88 | −1.503 [−2.237, −0.768] | <0.001 |
| FTD–HC | 52 | −1.473 [−2.368, −0.579] | 0.003 |
| AD–FTD | 59 | −0.373 [−1.168, 0.422] | 0.354 |

##### Posterior relative alpha (proportion)

Table S1b(v). Posterior relative-alpha contrasts.

| Contrast | n | Adjusted difference [95% HC4 CI] | Wild *p* |
| --- | --- | --- | --- |
| AD–HC | 65 | −0.039 [−0.058, −0.020] | <0.001 |
| AD+FTD–HC | 88 | −0.039 [−0.057, −0.020] | <0.001 |
| FTD–HC | 52 | −0.038 [−0.063, −0.013] | 0.004 |
| AD–FTD | 59 | −0.010 [−0.030, 0.010] | 0.343 |

Posterior absolute alpha uses O1/O2/P3/P4/Pz and summed 8–12-Hz linear power before nuisance adjustment. Global periodic expression uses all 15 non-frontopolar/non-lateral-frontal state channels. The PC1 score is not ordinary absolute power.

#### S1c. Peak detection and alpha-state quality

Table S1c. Individual alpha-peak detection and state-quality summaries.

| Group | IAF detected | Median pre-z alpha SD | Median split-half r | Median state condition number |
| --- | --- | --- | --- | --- |
| HC | 28/29 | 0.862 | 0.891 | 8.46 |
| AD | 20/36 | 0.830 | 0.898 | 6.85 |
| FTD | 19/23 | 0.842 | 0.896 | 7.38 |

IAF detection and quality are descriptive; no new quality threshold was used to exclude the primary cohort.

#### S1d. Spatial definitions and illustrative spectrum

Table S1d. Spatial measurement definitions and illustrative spectrum.

| Measurement | Construction |
| --- | --- |
| Theta / alpha / beta | 15-channel PC1: F3,F4,C3,C4,P3,P4,O1,O2,T3,T4,T5,T6,Fz,Cz,Pz; equal-participant average covariance over all 88 participants. Fp1, Fp2, F7, and F8 were omitted by a fixed policy because of their greater susceptibility to ocular and frontal artifact; Fp1/Fp2 were retained separately as nuisance references. |
| Gamma | 11-channel PC1: F3, F4, C3, C4, P3, P4, O1, O2, Fz, Cz, Pz; T3–T6 excluded from gamma covariance. Temporal electrodes remain in the other-band and aperiodic measurements. |
| Aperiodic state | Uniform mean exponent across the 15-channel set. |
| Frontal nuisance adjustment | Mean-centered Fp1/Fp2 reference regression at each 30–44-Hz bin, applied to F3/F4/Fz before decomposition; pooled-window mean preserved. |
| Regional alpha | HC-reference covariance (29 controls) for Anterior6 and Posterior7 bases; not the all-participant global basis. |
| Figure 2 example | sub-049 (HC), Pz; Window index: 99. The signed alpha-band mean is below the fitted background; this is a decomposition example, not a typical-alpha selection. The window index is stated in Figure 2. |

### Table S2. Transition order and adequacy

#### S2a. BIC preference on common VAR(3)-eligible target rows

Table S2a. Preferred VAR order on the common VAR(3)-eligible sample.

| Group | VAR(1) | VAR(2) | VAR(3) |
| --- | --- | --- | --- |
| All | 86 (97.7%) | 2 (2.3%) | 0 (0.0%) |
| HC | 29 (100.0%) | 0 (0.0%) | 0 (0.0%) |
| AD | 35 (97.2%) | 1 (2.8%) | 0 (0.0%) |
| FTD | 22 (95.7%) | 1 (4.3%) | 0 (0.0%) |

All three candidate orders use identical target rows within each participant. AIC, BIC, log likelihood and observation counts are retained in the source table.

#### S2b. Full-record VAR(1) diagnostics

Table S2b. Alpha residual-memory and stability diagnostics.

| Group | n | Raw alpha residual-memory failures | Detrended failures | Raw radius median / maximum | Detrended radius median / maximum |
| --- | --- | --- | --- | --- | --- |
| HC | 29 | 18/29 (62.1%) | 8/29 (27.6%) | 0.252 / 0.484 | 0.157 / 0.364 |
| AD | 36 | 7/36 (19.4%) | 6/36 (16.7%) | 0.228 / 0.615 | 0.162 / 0.463 |
| FTD | 23 | 4/23 (17.4%) | 5/23 (21.7%) | 0.195 / 0.475 | 0.136 / 0.437 |

Ljung–Box tests used 10 lags and a nominal 0.05 diagnostic threshold. These checks do not imply fully white innovations, exact first-order dynamics, or clinical inference. Full-record coefficients were not used for cross-validated predictions.

### Table S3. Selected source-ablation sensitivities and scale context

Coefficient values in S3a–S3c are in 10³Gα units. M0 adjusts for age and sex. Coefficient intervals are HC4 Wald 95% intervals; diagnosis p-values use the restricted wild bootstrap. S3a–S3b reproduce the six specifications in Figure 3C: the primary reference and five selected sensitivity analyses. The primary endpoint is unchanged; the sensitivity endpoints are separately re-estimated.

#### S3a. Selected specification estimates: AD–HC

Table S3a. AD–HC effects for the six specifications displayed in Figure 3C.

| Variant | n | Adjusted difference [95% CI] | Wild *p* |
| --- | --- | --- | --- |
| Primary VAR(1) | 65 | −3.190 [−5.254, −1.126] | 0.004 |
| Run-wise detrended | 65 | −1.915 [−3.504, −0.326] | 0.014 |
| VAR(2) | 65 | −3.010 [−5.595, −0.426] | 0.020 |
| VAR(3) | 65 | −2.228 [−4.528, 0.073] | 0.052 |
| 5-s boundary embargo | 65 | −3.337 [−5.392, −1.282] | 0.002 |
| IAF-relative (peak-detected subset) | 48 | −4.434 [−7.207, −1.661] | 0.003 |

#### S3b. Selected specification estimates: AD+FTD–HC

Table S3b. Pooled AD+FTD–HC effects for the six displayed specifications.

| Variant | n | Adjusted difference [95% CI] | Wild *p* |
| --- | --- | --- | --- |
| Primary VAR(1) | 88 | −2.857 [−4.892, −0.823] | 0.005 |
| Run-wise detrended | 88 | −1.611 [−3.038, −0.183] | 0.026 |
| VAR(2) | 88 | −2.340 [−4.875, 0.195] | 0.069 |
| VAR(3) | 88 | −1.652 [−3.884, 0.580] | 0.143 |
| 5-s boundary embargo | 88 | −2.966 [−4.995, −0.937] | 0.003 |
| IAF-relative (peak-detected subset) | 67 | −3.678 [−6.220, −1.136] | 0.004 |

These selected comparisons cover slow trends, temporal history, train/test proximity, and band definition. They do not constitute an exhaustive inventory of computed variants. Additional numerical outputs remain archived; the selected estimates do not establish stability across recording time or vigilance conditions.

Run-wise detrending removes a linear trend separately within each uninterrupted recording run, followed by state re-standardization and endpoint re-estimation. VAR(2) and VAR(3) include two and three one-second lags, respectively, and omit alpha at all included lags in the reduced model. The five-second embargo removes training pairs sharing observations with the held-out fold and training target windows within five seconds of a held-out target window in the same segment. All six specifications retain the primary five-state definition, including the 11-channel gamma spatial basis.

##### IAF-relative eligibility and interpretation

IAF is the most prominent qualifying peak in the median posterior periodic spectrum (O1, O2, P3, P4, Pz, T5, and T6) within 6–14 Hz, with prominence ≥0.05 log10 units. No IAF is imputed when a qualifying peak is absent. The alpha band is IAF±2 Hz; theta extends from 4 Hz to IAF−2 Hz and beta from IAF+2 Hz to 30 Hz. Gamma retains 30–44 Hz and the primary 11-channel basis.

The IAF-relative comparison includes 20 AD and 28 HC participants (n = 48), with 19 FTD participants additionally included in the pooled comparison (n = 67). These eligibility counts define the IAF-relative row rather than an additional sensitivity analysis. Its sample differs from that of the full-cohort primary reference. Without a separately displayed same-sample fixed-band comparison, differences in magnitude cannot distinguish band recentering from participant selection.

#### S3c. Primary breadth and target-specific estimates

Table S3c. Diagnostic breadth and target-specific primary effects.

| Endpoint | Contrast | n | Adjusted difference [95% CI] | Wild *p* |
| --- | --- | --- | --- | --- |
| Overall Gα | FTD–HC | 52 | −1.967 [−4.474, 0.540] | 0.121 |
| Overall Gα | AD–FTD | 59 | −1.428 [−3.126, 0.270] | 0.096 |
| α→θ | AD–HC | 65 | −1.984 [−4.309, 0.340] | 0.087 |
| α→β | AD–HC | 65 | −2.900 [−5.594, −0.206] | 0.032 |
| α→γ | AD–HC | 65 | −4.685 [−8.051, −1.320] | 0.006 |

Target coefficients are descriptive; compare targets using the direct clustered test in S6, not their separate p-values.

#### S3d. Lag support and denominator eligibility

Table S3d. Lag support, mean-only prediction comparison, and relative-share eligibility.

| Group | Valid lag pairs: median [IQR] / range | Relative-share eligible | MSE improvement: median [IQR] | Signed alpha share: median [IQR] |
| --- | --- | --- | --- | --- |
| HC | 804 [771, 877] range 701–963 | 19/29 | 0.003683 [-0.001908, 0.015851] | 0.155 [-0.007, 0.425] |
| AD | 803.5 [726.5, 868.25] range 303–1266 | 12/36 | -0.004545 [-0.006667, 0.003282] | -0.034 [-0.304, 0.101] |
| FTD | 690 [603, 816] range 478–997 | 7/23 | -0.002420 [-0.006658, 0.003556] | 0.265 [-0.149, 0.434] |

MSE improvement is the full model’s improvement over the training-fold mean-only predictor. The ratio is defined only above 10⁻¹² and may be negative or exceed one. Eligibility differs by diagnosis; medians are not population-wide proportions of neural information.

A positive denominator means only that the full model had lower average held-out MSE than the training-mean predictor; it is not a significance or performance-quality threshold. The full model includes target self-history as well as other states. Consequently, this denominator is not a measure of purely cross-rhythm predictability.

### Table S4. Expression/quality adjustment of Gα

M0 is age/sex-adjusted; M1 adds expression; MQ adds pre-z dynamic range and Fisher-z split-half reliability; M2 includes both; M2b additionally adjusts log state condition number. Rows are coefficient estimates in 10³ units. Retention is 100×adjusted/baseline, with diagnosis-stratified paired-bootstrap intervals. Negative or >100% interval limits are retained. “Withheld” means that the baseline sign/stability rule did not support a ratio interval, not that the model was absent.

## AD–HC

Table S4a(i). AD–HC contribution effects after expression/quality adjustment.

| Model | Expression | Diagnosis effect [95% HC4 CI] | Wild *p* | Retention % [95% bootstrap interval] |
| --- | --- | --- | --- | --- |
| M0 | — | −3.190 [−5.254, −1.126] | 0.004 | 100 (reference) |
| MQ | — | −2.147 [−4.237, −0.056] | 0.039 | 67.3 [16.9, 98.5] |
| M1 | Global periodic | −3.026 [−5.511, −0.541] | 0.019 | 94.9 [44.0, 131.2] |
| M2 | Global periodic | −2.419 [−4.633, −0.205] | 0.024 | 75.8 [18.6, 120.8] |
| M1 | Posterior periodic | −2.790 [−5.321, −0.259] | 0.031 | 87.5 [32.7, 128.5] |
| M2 | Posterior periodic | −2.376 [−4.761, 0.009] | 0.035 | 74.5 [16.2, 131.0] |
| M1 | Posterior relative | −2.551 [−4.992, −0.109] | 0.040 | 80.0 [12.2, 122.8] |
| M2 | Posterior relative | −2.200 [−4.536, 0.135] | 0.046 | 69.0 [6.3, 119.7] |
| M2b | Global periodic | −1.619 [−4.043, 0.806] | 0.163 | 50.7 [−35.8, 106.7] |

M0 is the age/sex-adjusted primary estimate for the same diagnostic contrast.

#### AD+FTD–HC

Table S4a(ii). Pooled AD+FTD–HC contribution effects after adjustment.

| Model | Expression | Diagnosis effect [95% HC4 CI] | Wild *p* | Retention % [95% bootstrap interval] |
| --- | --- | --- | --- | --- |
| M0 | — | −2.857 [−4.892, −0.823] | 0.005 | 100 (reference) |
| MQ | — | −1.538 [−3.512, 0.436] | 0.113 | 53.8 [−23.2, 85.2] |
| M1 | Global periodic | −2.523 [−4.833, −0.213] | 0.032 | 88.3 [29.6, 120.6] |
| M2 | Global periodic | −1.869 [−3.908, 0.170] | 0.059 | 65.4 [1.2, 108.8] |
| M1 | Posterior periodic | −2.213 [−4.504, 0.078] | 0.054 | 77.4 [4.4, 110.0] |
| M2 | Posterior periodic | −1.676 [−3.793, 0.440] | 0.103 | 58.7 [−17.2, 102.9] |
| M1 | Posterior relative | −1.961 [−4.166, 0.244] | 0.075 | 68.6 [−11.0, 105.7] |
| M2 | Posterior relative | −1.441 [−3.577, 0.696] | 0.153 | 50.4 [−38.2, 96.4] |
| M2b | Global periodic | −1.188 [−3.311, 0.934] | 0.242 | 41.6 [−49.1, 89.7] |

M0 is the age/sex-adjusted primary estimate for the same diagnostic contrast.

#### FTD–HC

Table S4a(iii). FTD–HC contribution effects after adjustment.

| Model | Expression | Diagnosis effect [95% HC4 CI] | Wild *p* | Retention % [95% bootstrap interval] |
| --- | --- | --- | --- | --- |
| M0 | — | −1.967 [−4.474, 0.540] | 0.121 | 100 (reference) |
| MQ | — | 0.166 [−2.150, 2.483] | 0.882 | -8.5; interval withheld |
| M1 | Global periodic | −1.538 [−4.389, 1.312] | 0.283 | 78.2; interval withheld |
| M2 | Global periodic | −0.287 [−2.707, 2.133] | 0.802 | 14.6; interval withheld |
| M1 | Posterior periodic | −1.250 [−4.053, 1.553] | 0.373 | 63.6; interval withheld |
| M2 | Posterior periodic | 0.010 [−2.575, 2.595] | 0.994 | -0.5; interval withheld |
| M1 | Posterior relative | −0.970 [−3.623, 1.684] | 0.462 | 49.3; interval withheld |
| M2 | Posterior relative | 0.243 [−2.415, 2.901] | 0.833 | -12.4; interval withheld |
| M2b | Global periodic | 0.764 [−1.410, 2.937] | 0.464 | -38.8; interval withheld |

M0 is the age/sex-adjusted primary estimate for the same diagnostic contrast.

#### S4b. Paired coefficient changes and interaction diagnostic

Table S4b. AD–HC contribution coefficient changes relative to M0.

| Model | Adjusted minus M0 [95% bootstrap interval] | Valid draws | Ratio stable |
| --- | --- | --- | --- |
| MQ | 1.043 [0.049, 2.153] | 5000 | Yes |
| M1 | 0.164 [−0.962, 1.264] | 5000 | Yes |
| M2 | 0.771 [−0.537, 2.137] | 5000 | Yes |

AD–HC, M3 diagnosis × expression: −0.589 [−3.084, 1.905]; HC4 p = 0.65. This interaction is not the M2 diagnosis effect.

AD+FTD–HC, M3 diagnosis × expression: −0.055 [−2.174, 2.064]; HC4 p = 0.96. This interaction is not the M2 diagnosis effect.

FTD–HC, M3 diagnosis × expression: 1.489 [−1.259, 4.237]; HC4 p = 0.29. This interaction is not the M2 diagnosis effect.

The coefficient-change interval remains informative when a ratio is poorly identified. No formal cross-endpoint retention-difference test is reported.

### Table S5. Conditional alpha self-persistence

M0 is age/sex-adjusted; M1 adds expression; MQ adds pre-z dynamic range and Fisher-z split-half reliability; M2 includes both; M2b additionally adjusts log state condition number. Rows are coefficient estimates in 10³ units. Retention is 100×adjusted/baseline, with diagnosis-stratified paired-bootstrap intervals. Negative or >100% interval limits are retained. “Withheld” means that the baseline sign/stability rule did not support a ratio interval, not that the model was absent.

## AD–HC

Table S5a. AD–HC conditional alpha self-persistence after adjustment.

| Model | Expression | Diagnosis effect [95% HC4 CI] | Wild *p* | Retention % [95% bootstrap interval] |
| --- | --- | --- | --- | --- |
| M0 | — | −73.863 [−119.738, −27.988] | 0.003 | 100 (reference) |
| MQ | — | −2.783 [−29.177, 23.612] | 0.823 | 3.8 [−49.2, 34.8] |
| M1 | Global periodic | −23.772 [−74.184, 26.640] | 0.351 | 32.2 [−60.9, 76.1] |
| M2 | Global periodic | −3.283 [−31.595, 25.030] | 0.814 | 4.4 [−52.6, 36.8] |
| M1 | Posterior periodic | −14.744 [−62.200, 32.711] | 0.533 | 20.0 [−76.6, 63.0] |
| M2 | Posterior periodic | −0.181 [−29.689, 29.328] | 0.996 | 0.2 [−67.1, 34.7] |
| M1 | Posterior relative | −6.980 [−54.055, 40.094] | 0.765 | 9.5 [−96.9, 54.7] |
| M2 | Posterior relative | 4.655 [−23.908, 33.218] | 0.747 | −6.3 [−77.8, 27.2] |
| M2b | Global periodic | −2.122 [−33.383, 29.138] | 0.898 | 2.9 [−57.0, 41.6] |

Pα is an in-sample conditional coefficient; its comparison with out-of-sample Gα is not a formal matched negative control.

#### S5b. Paired coefficient changes and interaction diagnostic

Table S5b. Self-persistence coefficient changes relative to M0.

| Model | Adjusted minus M0 [95% bootstrap interval] | Valid draws | Ratio stable |
| --- | --- | --- | --- |
| MQ | 71.081 [29.902, 111.131] | 5000 | Yes |
| M1 | 50.091 [16.056, 91.671] | 5000 | Yes |
| M2 | 70.581 [30.781, 111.307] | 5000 | Yes |

AD–HC, M3 diagnosis × expression: −0.924 [−39.484, 37.637]; HC4 p = 0.96. This interaction is not the M2 diagnosis effect.

The coefficient-change interval remains informative when a ratio is poorly identified. No formal cross-endpoint retention-difference test is reported.

### Table S6. Source, target and regional profiles

All effects below are in 10³ predictive-contribution units. CR1 participant-clustered covariance and cluster t-based intervals differ from HC4 intervals in S4–S5. M2 applies alpha-expression and alpha-quality covariates to every source. These exploratory comparisons have no multiplicity adjustment.

Table S6a(i). Source-specific AD–HC effects.

| Model | Component | AD–HC difference [95% clustered CI] | Descriptive *p* |
| --- | --- | --- | --- |
| M0 | theta | 0.500 [−0.496, 1.497] | 0.323 |
| M0 | alpha | −3.190 [−5.302, −1.078] | 0.004 |
| M0 | beta | −1.598 [−3.350, 0.154] | 0.073 |
| M0 | gamma | −0.691 [−2.168, 0.786] | 0.355 |
| M2 | theta | 0.079 [−0.914, 1.073] | 0.876 |
| M2 | alpha | −2.419 [−4.656, −0.182] | 0.035 |
| M2 | beta | −1.544 [−3.057, −0.030] | 0.046 |
| M2 | gamma | −0.649 [−2.116, 0.818] | 0.385 |

Table S6a(ii). Diagnosis-by-source omnibus and direct alpha-specificity contrasts.

| Model | Omnibus | Omnibus *p* | α−mean others | Direct *p* |
| --- | --- | --- | --- | --- |
| M0 | F(3,64) = 5.047 | 0.0034 | −2.594 [−4.560, −0.628] | 0.011 |
| M2 | F(3,64) = 2.836 | 0.045 | −1.714 [−4.002, 0.573] | 0.145 |

Table S6b(i). Alpha contributions to individual future targets.

| Model | Component | AD–HC difference [95% clustered CI] | Descriptive *p* |
| --- | --- | --- | --- |
| M0 | theta | −1.984 [−4.355, 0.386] | 0.099 |
| M0 | beta | −2.900 [−5.660, −0.140] | 0.040 |
| M0 | gamma | −4.685 [−8.135, −1.235] | 0.009 |
| M2 | theta | −1.755 [−4.385, 0.875] | 0.195 |
| M2 | beta | −3.477 [−6.582, −0.372] | 0.029 |
| M2 | gamma | −2.024 [−5.127, 1.079] | 0.204 |

Table S6b(ii). Diagnosis-by-target omnibus and beta–gamma contrasts.

| Model | Omnibus | Omnibus *p* | β−γ contrast | Direct *p* |
| --- | --- | --- | --- | --- |
| M0 | F(2,64) = 1.093 | 0.34 | 1.785 [−1.830, 5.400] | 0.333 |
| M2 | F(2,64) = 0.522 | 0.60 | −1.453 [−4.710, 1.803] | 0.385 |

Table S6c. Regional alpha-source effects.

| Region | M2 AD–HC difference [95% clustered CI] | Descriptive *p* |
| --- | --- | --- |
| Anterior6 | −1.195 [−2.466, 0.076] | 0.065 |
| Posterior7 | −2.326 [−4.752, 0.100] | 0.060 |

Posterior7 minus anterior6 diagnosis difference: −1.131 [−3.477, 1.215]; p = 0.34.

HC-reference bases (29 controls); region-specific nuisance slopes. Non-significance does not establish equivalence.

### Table S7. Complementarity and supporting analyses

#### S7a. Joint displacement and reciprocal model comparisons

AD–HC joint test: F(2,64) = 9.523, p < 0.001; n = 65. Diagnosis coefficients were 0.757 for D_E and 0.628 for D_G. Clustered covariance entries were 0.060214 (EE), −0.002683 (EG), and 0.046171 (GG). This tests joint displacement; the reciprocal comparisons below test complementarity.

Table S7a. Reciprocal nested logistic-model comparisons.

| Nested comparison | df | LR statistic | Raw *p* |
| --- | --- | --- | --- |
| Contribution added to expression model | 1 | 6.7285 | 0.010 |
| Expression added to contribution model | 1 | 7.4862 | 0.006 |
| Expression added to demographics | 1 | 9.4853 | 0.002 |
| Contribution added to demographics | 1 | 8.7276 | 0.003 |

The first two rows are the reciprocal complementarity tests. Applying Holm correction within this two-test family gives adjusted p = 0.012 for both. The two demographic-only comparisons are reported nominally and are not included in that family.

#### S7b. Unadjusted effect sizes and correlations

Table S7b(i). Standardized group differences in the two components and composite.

| Endpoint | Cohen’s d [95% bootstrap interval] | HC / AD n |
| --- | --- | --- |
| Alpha-expression deficit | 0.890 [0.405, 1.476] | 29 / 36 |
| Alpha predictive-contribution deficit | 0.781 [0.331, 1.262] | 29 / 36 |
| Dual-alpha score | 1.151 [0.648, 1.777] | 29 / 36 |

Table S7b(ii). Correlations between expression and contribution deficits.

| Correlation sample | r [95% bootstrap interval] | n |
| --- | --- | --- |
| HC | 0.070 [−0.290, 0.422] | 29 |
| HC + AD | 0.201 [−0.046, 0.449] | 65 |

Reciprocal logistic comparisons, not low correlation, support incremental group information. Effect-size intervals do not constitute a direct superiority test.

#### S7c. Half-recording reliability and illustrative projections

Table S7c. Half-recording reliability and illustrative full-record projections.

| Endpoint | ICC(2,1) [95% interval] | n | SB projected full-recording ICC | Undefined projection fraction |
| --- | --- | --- | --- | --- |
| Alpha expression | 0.922 [0.876, 0.953] | 88 | 0.959 | 0.000 |
| Gα | 0.208 [−0.124, 0.419] | 88 | 0.344 | 0.112 |
| Dual-alpha score | 0.429 [0.191, 0.610] | 88 | 0.600 | 0.000 |

Spearman–Brown projections assume parallel segments with independent errors. They are not validated duration recommendations. The Gα ICC interval spans zero, making extrapolation especially uncertain.

#### S7d. Apparent AUC and paired AUC differences

Table S7d(i). Apparent within-cohort discrimination.

| Score | AUC [95% bootstrap interval] | n |
| --- | --- | --- |
| Alpha expression | 0.747 [0.625, 0.861] | 65 |
| Gα | 0.666 [0.522, 0.797] | 65 |
| Dual-alpha score | 0.799 [0.679, 0.909] | 65 |

Table S7d(ii). Paired differences in apparent AUC.

| Paired comparison | ΔAUC [95% interval] | Bootstrap *p* |
| --- | --- | --- |
| Dual − expression | 0.052 [−0.024, 0.134] | 0.194 |
| Dual − contribution | 0.133 [0.016, 0.267] | 0.026 |

These are apparent within-cohort score-discrimination summaries. No external diagnostic validation, decision threshold or prospective classifier is established.

#### S7e. MMSE associations

Table S7e. Exploratory associations with MMSE.

| Sample | Model | Term | n | MMSE difference per SD [95% HC4 CI] | Wild *p* |
| --- | --- | --- | --- | --- | --- |
| AD+FTD | E only | D_E | 59 | 0.234 [−0.784, 1.252] | 0.664 |
| AD+FTD | Gα only | D_G | 59 | 0.379 [−0.551, 1.309] | 0.435 |
| AD+FTD | Dual | Dual score | 59 | 0.376 [−0.438, 1.189] | 0.397 |
| AD+FTD | E + Gα | D_E | 59 | 0.186 [−0.955, 1.326] | 0.753 |
| AD+FTD | E + Gα | D_G | 59 | 0.351 [−0.680, 1.383] | 0.485 |
| AD | E only | D_E | 36 | −0.016 [−1.614, 1.583] | 0.990 |
| AD | Gα only | D_G | 36 | 0.411 [−1.061, 1.883] | 0.585 |
| AD | Dual | Dual score | 36 | 0.188 [−1.566, 1.941] | 0.763 |
| AD | E + Gα | D_E | 36 | −0.046 [−1.915, 1.822] | 0.965 |
| AD | E + Gα | D_G | 36 | 0.415 [−1.383, 2.213] | 0.607 |

All reported associations are exploratory. Null associations are retained; HC participants are not used to infer severity from their constant MMSE scores.

#### S7f. Logistic coefficient summaries

Table S7f. Logistic-model coefficient estimates.

| Model | Term | Log-odds coefficient [95% HC3 CI] | HC3 *p* |
| --- | --- | --- | --- |
| Demographics | age | −0.246 [−0.734, 0.241] | 0.324 |
| Demographics | male | −0.601 [−1.115, −0.087] | 0.022 |
| Expression | age | −0.303 [−0.820, 0.215] | 0.255 |
| Expression | male | −0.496 [−1.045, 0.054] | 0.077 |
| Expression | D_E | 0.933 [0.294, 1.573] | 0.004 |
| Contribution | age | −0.301 [−0.792, 0.191] | 0.233 |
| Contribution | male | −0.565 [−1.119, −0.011] | 0.046 |
| Contribution | D_G | 0.940 [0.323, 1.556] | 0.003 |
| Joint | age | −0.331 [−0.841, 0.179] | 0.204 |
| Joint | male | −0.473 [−1.056, 0.111] | 0.117 |
| Joint | D_E | 0.867 [0.191, 1.543] | 0.012 |
| Joint | D_G | 0.889 [0.130, 1.647] | 0.022 |

HC3 coefficient inference is distinct from the likelihood-ratio tests in S7a. Predictors were standardized within the HC–AD logistic-analysis sample.

### Table S8. Exploratory circuit methods and results

#### S8a. Model and evaluation specification

Each JR column contains excitatory, pyramidal and inhibitory population modules. With V = y₁ − y₂ and S(V) = 5/[1 + exp{0.56(6 − V)}], the local synaptic equations are:

$$\ddot{y}_{0}=H_{e}aS\left( y_{1}-y_{2} \right)-2a\dot{y}_{0}-a^{2}y_{0}$$

$$\ddot{y}_{1}=H_{e}a\left[ p\left( t \right)+C_{2}S\left( C_{1}y_{0} \right) \right]-2a\dot{y}_{1}-a^{2}y_{1}$$

$$\ddot{y}_{2}=H_{i}bC_{4}S\left( C_{3}y_{0} \right)-2b\dot{y}_{2}-b^{2}y_{2}$$

The observation is V, not any synaptic parameter. Rate constants and gains are jointly multiplied by q for each downstream column. The directed source-derived input is K·S[Vα(t − 20 ms)]. Reducing K lowers both tonic and fluctuating input; it is not a selective perturbation of fluctuating information alone.

Table S8a(i). Circuit parameters and evaluation specification.

| Module | Setting |
| --- | --- |
| Reference local JR | Hₑ = 3.25 mV; Hᵢ = 22 mV; a = 100 s⁻¹; b = 50 s⁻¹; C₁ = 135; C₂ = 108; C₃ = C₄ = 33.75 |
| Kinetic factors | q = 1, 0.55, 1.8, 3.0; a, b, Hₑ and Hᵢ each multiplied by q |
| OU inputs | Independent per column; means 100,90,90,90 s⁻¹; SD 22 s⁻¹; correlation time 500 ms |
| Interventions | Source C₂: 108→105.84; common downstream-input K: 10→5; both applied together |
| Observation | Equal weights 0.25 per column; 200-Hz sampling; 0.5–45-Hz fourth-order zero-phase Butterworth filter |
| Integration / duration | Fourth-order Runge–Kutta; 1-ms step; 20-s burn-in; 24 paired 600-s retained realizations |
| Evaluation seed / bootstrap | 2026092301 (not used in preliminary simulations); 5,000 paired-realization percentile-bootstrap resamples |
| Grid / assumptions | 25 grid cells, 4 paired 300-s realizations per cell; 11 assumption settings, 8 paired 400-s realizations per setting |
| Measurement | Same one-second spectral fitting and blocked-ablation definitions; no human scalp-channel spatial PC1 in the model observation |

The configuration was selected in preliminary simulations and then held fixed for evaluation seed 2026092301, a seed not used in those simulations. Controls change only the operation listed below, using the same stochastic realization as their reference. No patient-level parameter fitting is claimed.

Table S8a(ii). Control interventions and their interpretation.

| Control | Operation relative to reference | Question addressed |
| --- | --- | --- |
| Mean downstream input restored | Set K = 5 and add (10 − 5) times the post-burn-in reference source mean firing rate to each downstream input. Local source recurrence and observation weights are unchanged. | Separates loss of mean recipient drive from reduced fluctuating source-derived input. |
| Source observation attenuated | Multiply only the source observation weight by 0.7 (0.25 → 0.175); leave each downstream weight at 0.25. Do not renormalize. Neural dynamics and K remain unchanged. | Tests dependence on the voltage mixture rather than a neural intervention. |
| Source input shifted by 30 s | Circularly shift the simulated source firing-rate sequence by 30 s before applying the 20-ms delay and K = 10 downstream. The unshifted source voltage remains in the observation. | A nonphysiological temporal-alignment surrogate, not a physiological conduction delay. |
| Downstream input removed | Set K = 0; retain independent downstream OU drives, source recurrence and observation weights. | Removes the source-derived projection; does not guarantee a zero spectral-ablation statistic. |

#### S8b. Condition means

Table S8b. Circuit condition means and 95% percentile-bootstrap intervals.

| Condition (24 realizations) | Periodic expression, dB [95% interval] | 10³Gα [95% interval] |
| --- | --- | --- |
| Reference | 9.519 [9.044, 10.036] | 8.369 [3.766, 12.288] |
| Local recurrence reduced | 6.492 [5.541, 7.429] | 14.527 [11.150, 17.999] |
| Downstream input reduced | 9.045 [8.581, 9.520] | −1.440 [−3.809, 0.668] |
| Both reduced | 6.190 [5.351, 7.037] | −2.328 [−6.297, 0.965] |
| Downstream input removed | 9.410 [8.968, 9.831] | 2.959 [−0.571, 7.089] |
| Reduced input; mean restored | 9.650 [9.144, 10.162] | 4.724 [2.022, 7.510] |
| Source observation attenuated | 7.944 [7.530, 8.351] | 7.970 [3.908, 11.667] |
| Source input shifted 30 s | 9.390 [8.902, 9.889] | 1.734 [−0.060, 3.632] |

These are condition-mean intervals, not paired-change intervals. Negative Gα is permitted by held-out refitting.

#### S8c. All planned paired changes

##### Periodic expression (dB)

Table S8c(i). Paired changes in periodic-alpha expression.

| Condition minus paired reference | Change [95% paired bootstrap interval] |
| --- | --- |
| Both reduced | −3.329 [−4.413, −2.215] |
| Local recurrence reduced | −3.028 [−4.188, −1.834] |
| Reduced input; mean restored | 0.131 [0.050, 0.208] |
| Source observation attenuated | −1.575 [−1.646, −1.500] |
| Downstream input reduced | −0.474 [−0.607, −0.327] |
| Downstream input removed | −0.109 [−0.237, 0.025] |
| Source input shifted 30 s | −0.129 [−0.236, −0.017] |

##### Predictive contribution (10³Gα)

Table S8c(ii). Paired changes in alpha predictive contribution.

| Condition minus paired reference | Change [95% paired bootstrap interval] |
| --- | --- |
| Both reduced | −10.697 [−17.569, −4.134] |
| Local recurrence reduced | 6.158 [−0.084, 12.559] |
| Reduced input; mean restored | −3.645 [−7.191, −0.150] |
| Source observation attenuated | −0.399 [−1.203, 0.421] |
| Downstream input reduced | −9.809 [−14.826, −4.860] |
| Downstream input removed | −5.410 [−10.526, −0.442] |
| Source input shifted 30 s | −6.634 [−10.175, −2.669] |

##### Conventional alpha power (dB)

Table S8c(iii). Paired changes in conventional alpha power.

| Condition minus paired reference | Change [95% paired bootstrap interval] |
| --- | --- |
| Both reduced | −0.953 [−1.263, −0.651] |
| Local recurrence reduced | −0.983 [−1.243, −0.722] |
| Reduced input; mean restored | −0.184 [−0.230, −0.135] |
| Source observation attenuated | −2.645 [−2.683, −2.604] |
| Downstream input reduced | 0.079 [−0.001, 0.151] |
| Downstream input removed | 0.235 [0.097, 0.373] |
| Source input shifted 30 s | −0.164 [−0.226, −0.097] |

##### Uncorrected-band contribution (10³G)

Table S8c(iv). Paired changes in uncorrected-band contribution.

| Condition minus paired reference | Change [95% paired bootstrap interval] |
| --- | --- |
| Both reduced | −1.304 [−6.099, 3.647] |
| Local recurrence reduced | 1.343 [−3.759, 6.348] |
| Reduced input; mean restored | −0.418 [−2.707, 1.777] |
| Source observation attenuated | −1.832 [−2.684, −0.908] |
| Downstream input reduced | −2.017 [−4.754, 0.870] |
| Downstream input removed | −0.984 [−5.029, 3.199] |
| Source input shifted 30 s | −3.222 [−5.715, −0.725] |

##### Conditional alpha self-persistence

Table S8c(v). Paired changes in conditional alpha self-persistence.

| Condition minus paired reference | Change [95% paired bootstrap interval] |
| --- | --- |
| Both reduced | 0.022 [−0.006, 0.047] |
| Local recurrence reduced | −0.014 [−0.045, 0.013] |
| Reduced input; mean restored | 0.024 [0.010, 0.037] |
| Source observation attenuated | −0.038 [−0.043, −0.034] |
| Downstream input reduced | 0.005 [−0.012, 0.022] |
| Downstream input removed | −0.007 [−0.025, 0.013] |
| Source input shifted 30 s | 0.016 [0.003, 0.029] |

Paired changes use the same stochastic realization across conditions. Restoring mean input substantially attenuated the contribution contrast in the reported evaluation. The interval should not be taken as evidence of platform-invariant incomplete restoration. Local recurrent-excitation reduction can also change spectral regime. These are conditional model findings, not identification of an anatomical or disease mechanism.

#### S8d. Human–model measurement check

Table S8d(i). Human and model measurement distributions.

| Group | Measure | n | Mean [95% percentile-bootstrap interval] |
| --- | --- | --- | --- |
| Controls | Expression (dB) | 29 | 3.447 [2.991, 3.928] |
| Controls | Self-persistence | 29 | 0.140 [0.107, 0.175] |
| Controls | 10³Gα | 29 | 2.191 [0.436, 4.089] |
| Model | Expression (dB) | 24 | 9.519 [9.026, 10.017] |
| Model | Self-persistence | 24 | 0.468 [0.445, 0.487] |
| Model | 10³Gα | 24 | 8.369 [3.816, 12.306] |

Table S8d(ii). Model values inside the observed HC range.

| Measure | Model values inside observed HC range | HC range (native units) |
| --- | --- | --- |
| Expression (dB) | 0/24 (0.0%) | 1.33886 to 6.59608 |
| Self-persistence | 1/24 (4.2%) | 0.00574 to 0.40935 |
| Gα | 15/24 (62.5%) | −0.00421 to 0.01535 |

The separate measurement-check bootstrap has the same model means but may have slightly different Monte Carlo interval limits from S8b. Human expression averages 15 channels; human dynamic states use spatial PC1, including gamma11; simulation values use one equal-weight voltage mixture. Range overlap is descriptive, not a model-validity test.

#### S8e. Source spectral characteristics

Table S8e. Source-rhythm characteristics under each condition.

| Condition | Alpha-dominant source traces | Median dominant frequency (Hz) |
| --- | --- | --- |
| Reference | 24/24 | 10.25 |
| Local recurrence reduced | 13/24 | 10.00 |
| Downstream input reduced | 24/24 | 10.25 |
| Both reduced | 13/24 | 10.00 |
| Downstream input removed | 24/24 | 10.25 |
| Reduced input; mean restored | 24/24 | 10.25 |
| Source observation attenuated | 24/24 | 10.25 |
| Source input shifted 30 s | 24/24 | 10.25 |

Eleven of 24 local/combined source traces were no longer alpha-dominant, so the local expression change can include a change in spectral regime rather than a pure amplitude reduction.

#### S8f. Complete parameter-response grid

Table S8f. Complete recurrent-excitation/downstream-input parameter grid.

| Local multiplier ℓ | Input multiplier κ | Mean expression (dB) | Mean 10³Gα |
| --- | --- | --- | --- |
| 0.96 | 0.00 | 3.001 | 4.006 |
| 0.96 | 0.25 | 2.677 | −7.905 |
| 0.96 | 0.50 | 2.692 | 7.045 |
| 0.96 | 0.75 | 2.645 | 6.346 |
| 0.96 | 1.00 | 2.495 | 15.698 |
| 0.97 | 0.00 | 4.342 | 1.425 |
| 0.97 | 0.25 | 4.012 | −7.472 |
| 0.97 | 0.50 | 4.048 | 3.464 |
| 0.97 | 0.75 | 3.734 | 16.887 |
| 0.97 | 1.00 | 4.116 | 23.935 |
| 0.98 | 0.00 | 6.649 | −2.546 |
| 0.98 | 0.25 | 6.510 | −0.687 |
| 0.98 | 0.50 | 6.306 | 3.002 |
| 0.98 | 0.75 | 6.072 | −1.316 |
| 0.98 | 1.00 | 6.238 | 3.897 |
| 0.99 | 0.00 | 7.856 | 3.638 |
| 0.99 | 0.25 | 7.617 | −6.786 |
| 0.99 | 0.50 | 7.774 | −0.350 |
| 0.99 | 0.75 | 7.539 | 10.384 |
| 0.99 | 1.00 | 7.534 | 13.153 |
| 1.00 | 0.00 | 8.802 | 2.259 |
| 1.00 | 0.25 | 8.488 | −0.638 |
| 1.00 | 0.50 | 8.441 | −3.199 |
| 1.00 | 0.75 | 8.379 | −1.520 |
| 1.00 | 1.00 | 8.682 | 11.128 |

Each cell uses 4 paired 300-s realizations, not the 24-pair main evaluation. Every supplied cell is retained; no favorable cell is selected for reporting.

#### S8g. Operating-assumption analyses for the two alpha measurements

Table S8g. Paired effects under alternative operating assumptions.

| Assumption setting | Type | Δ expression, dB [95% interval] | Δ10³Gα [95% interval] |
| --- | --- | --- | --- |
| Reference settings (additional realizations) | Reduced recurrence | −3.008 [−4.683, −1.381] | 3.166 [−9.541, 17.991] |
| Reference settings (additional realizations) | Reduced input | −0.503 [−0.726, −0.292] | −7.663 [−17.198, 1.546] |
| Reference settings (additional realizations) | Both reduced | −3.182 [−4.758, −1.656] | −2.527 [−13.864, 9.991] |
| OU input correlation time: 150 ms | Reduced recurrence | −0.352 [−0.499, −0.201] | −5.094 [−11.323, 0.527] |
| OU input correlation time: 150 ms | Reduced input | 0.648 [0.537, 0.751] | −4.850 [−11.344, 1.560] |
| OU input correlation time: 150 ms | Both reduced | 0.389 [0.266, 0.525] | −4.121 [−9.433, 0.885] |
| OU input correlation time: 250 ms | Reduced recurrence | −1.472 [−2.079, −0.897] | −22.940 [−26.046, −20.194] |
| OU input correlation time: 250 ms | Reduced input | 0.233 [−0.052, 0.504] | −21.597 [−25.226, −17.881] |
| OU input correlation time: 250 ms | Both reduced | −0.970 [−1.624, −0.355] | −28.141 [−32.216, −23.592] |
| OU input correlation time: 750 ms | Reduced recurrence | −2.416 [−4.014, −1.095] | 3.289 [−5.205, 12.495] |
| OU input correlation time: 750 ms | Reduced input | −0.514 [−0.731, −0.279] | −4.565 [−14.363, 2.607] |
| OU input correlation time: 750 ms | Both reduced | −2.494 [−4.202, −1.133] | −7.796 [−14.469, −2.438] |
| Source-to-recipient delay: 0 ms | Reduced recurrence | −3.132 [−4.865, −1.462] | 14.623 [−2.215, 34.237] |
| Source-to-recipient delay: 0 ms | Reduced input | −0.666 [−0.969, −0.340] | −2.136 [−11.761, 12.647] |
| Source-to-recipient delay: 0 ms | Both reduced | −3.286 [−4.950, −1.656] | 3.010 [−5.649, 16.363] |
| Source-to-recipient delay: 40 ms | Reduced recurrence | −2.858 [−4.609, −1.097] | 9.642 [−5.236, 28.132] |
| Source-to-recipient delay: 40 ms | Reduced input | −0.134 [−0.555, 0.257] | −4.511 [−11.099, 5.786] |
| Source-to-recipient delay: 40 ms | Both reduced | −2.895 [−4.542, −1.281] | −5.240 [−14.978, 5.125] |
| Integration step: 0.5 ms | Reduced recurrence | −4.336 [−6.047, −2.461] | 7.036 [−4.767, 18.409] |
| Integration step: 0.5 ms | Reduced input | −0.556 [−0.881, −0.259] | −8.008 [−15.087, −0.955] |
| Integration step: 0.5 ms | Both reduced | −4.432 [−6.045, −2.631] | −6.887 [−18.118, 4.275] |
| Identical column kinetics (all q = 1) | Reduced recurrence | −2.725 [−4.791, −0.780] | 4.731 [−5.128, 13.838] |
| Identical column kinetics (all q = 1) | Reduced input | −3.286 [−4.146, −2.283] | 2.418 [−3.549, 8.112] |
| Identical column kinetics (all q = 1) | Both reduced | −4.821 [−6.436, −3.125] | 2.935 [−6.442, 13.207] |
| Observation: source only | Reduced recurrence | −4.963 [−8.049, −1.932] | −7.384 [−22.287, 3.357] |
| Observation: source only | Reduced input | 0.000 [0.000, 0.000] | 0.000 [0.000, 0.000] |
| Observation: source only | Both reduced | −4.963 [−8.040, −1.954] | −7.384 [−21.524, 3.438] |
| Observation: source weight 1/2 | Reduced recurrence | −3.899 [−6.190, −1.665] | 8.518 [−3.173, 20.591] |
| Observation: source weight 1/2 | Reduced input | −0.309 [−0.516, −0.118] | −7.176 [−15.749, 1.795] |
| Observation: source weight 1/2 | Both reduced | −3.948 [−6.122, −1.836] | 2.071 [−9.312, 18.503] |
| Observation: source weight 1/8 | Reduced recurrence | −2.063 [−3.190, −0.973] | 0.645 [−6.862, 8.668] |
| Observation: source weight 1/8 | Reduced input | −0.558 [−0.712, −0.408] | −4.455 [−8.603, −1.216] |
| Observation: source weight 1/8 | Both reduced | −2.300 [−3.299, −1.308] | −5.704 [−11.963, 1.240] |

Each operating-assumption setting uses eight paired 400-s realizations. All planned settings and both directions of change are retained; these checks delimit generalization beyond the chosen regime.

#### Software environment

The reported evaluation used macOS 15.7.9 (arm64), Python 3.12.13, NumPy 2.4.5, pandas 3.0.3, SciPy 1.17.1, statsmodels 0.14.6, h5py 3.12.1, Matplotlib 3.10.0, and Numba 0.65.1. Seed reproducibility does not imply bitwise cross-platform identity for nonlinear stochastic simulations. Model conclusions are restricted to the documented operating regime.
